# Investigation of sex- and age-specific effects of genetic risk for depression on peripheral biomarkers

**DOI:** 10.1101/2022.11.20.22282570

**Authors:** Earvin S. Tio, Jennifer S. Rabin, Liisa A. M. Galea, Daniel Felsky

## Abstract

**Background:** Polygenic risk for depression is associated with elevated white blood cell count (WBC), providing support for a pro-inflammatory causal mechanism of depression. However, the important roles of biological sex and age on depression and inflammation have not been accounted for in these analyses.

**Methods:** We calculated polygenic risk scores for depression (PRS_dep_) in 362,074 individuals from the UK Biobank (aged 39-72, 53.7% female) and 22,965 individuals from the Canadian Longitudinal Study on Aging (CLSA; aged 45-86, 50.3% female). We tested for main effects of PRS_dep_, sex, and non-linear age, as well as their interactions, on 25 blood-based measures (i.e., from complete blood counts, metabolic and lipid panels, and inflammatory tests). Associations significant in both cohorts were carried forward for further sex-stratified and mediation modelling.

**Results:** PRS_dep_ was significantly associated with 13 biomarkers in the UK Biobank, with five replicating in the CLSA (WBC, granulocyte, and lymphocyte counts, and C-reactive protein and triglyceride levels), with standardized effect sizes ranging from β=0.006 (C-reactive protein, *p*=7.50×10^−4^) to β=0.03 (WBC, *p*=1.71×10^−4^). Sex-specific effects of PRS_dep_ were observed for C-reactive protein levels (male; Stouffer’s *p*=2.40 ×10^−5^) and triglyceride levels (female; Stouffer’s *p*=5.19 ×10^−3^). Further differentiation was observed in the female subgroups based on post-menopausal status for WBC (Stouffer’s *p*=3.83 ×10^−3^) and neutrophil/granulocyte (Stouffer’s *p*=5.05 ×10^−3^) counts. Bi-directional mediation was also observed between all five biomarkers and a depression diagnosis, with up to 12.6% of the association between PRS_dep_ and triglyceride levels mediated through depression.

**Conclusions:** We have shown that effects of PRS_dep_ on multiple peripheral biomarkers, beyond WBC, remain significant even when accounting for important interactions of biological sex and age, providing insight into the pro-inflammatory etiology of depression.

## Introduction

An association between genetic risk for depression and white blood cell (WBC) count was recently established by the PsycheMERGE network, providing supporting evidence for a pro-inflammatory state in depression and promoting WBC count as a potential biomarker^1^. Elevated WBC count has also been associated with subsequent risk for psychiatric disorders in the Swedish Apolipoprotein Mortality Risk (AMORIS) cohort^2^. Furthermore, dysregulation of the immune system - specifically cell-mediated processes - has been shown to play a role in the etiology of major depressive disorder (MDD)^3,4^. In particular, levels of both pro- and anti-inflammatory cytokines such as tumor necrosis factor (TNF)α, interferon (IFN)γ, and certain members of the interleukin (IL) family, amongst others, have been associated with depression^5^. These cytokines cascade through immune-related pathways that increase leukocyte production and recruitment^6^. Beyond WBC, other putative biomarkers including total cholesterol, high-density lipoprotein, and triglyceride levels have also been associated with MDD via vascular aging and metabolic syndromes^7,8^. Lipid metabolism, in particular, is a key process in immune cell activation and modulation^9^.

Genetic studies of MDD also implicate immune-related risk variants that are linked to cytokine production^10,11^. However, as with most genetic studies of psychiatric phenotypes, the estimated effect sizes of these variants are small and inconsistent across studies^12^. Mendelian randomization analyses in the AMORIS cohort suggest a putative causal relationship between WBC and depression^2^. A recent genome-wide meta-analysis of both clinical and broadly-defined depression identified 44 independent risk variants pointing to a potential polygenic structure underlying the disorder^13^. However, despite well-powered genome-wide association studies (GWAS), polygenic risk scores (PRS) - which aggregate the effects of individual risk variants across the genome - have not yet shown clinical utility in isolation and may be best used as adjuncts to clinical evaluation. Nonetheless, PRSs can be effective tools in understanding the common and unique biological mechanisms across traits (e.g., depression and immune function) due to the inherently fixed nature of genetic variants across the lifespan. This genetic causality, however, can be over-estimated when the measured outcome is subject to environmental factors^14^. As such, models associating inflammatory markers with PRSs for depression must account for the influences of age, sex, and inflammatory conditions, which are all known to impact both leukocyte counts and depressive phenotypes^15^.

Lifespan analyses in community populations show that the relationship between inflammatory biomarkers and depressive symptoms vary by age. For example, the association between blood-derived measures of interleukin 6 (IL-6) and C-reactive protein (CRP) and anhedonia decreases with age, whereas the association with somatic complaints increases, with a significant sex effect in the latter association^16^. Furthermore, the prevalence and multifactorial etiology of depression are known to vary across the adult lifespan. For example, aging-associated vascular dysfunction can contribute to both cardiovascular and cerebrovascular related depressive symptomatology that manifest later in life^17^. Phenotypic variations such as late-life depression (LLD; defined as depression in those aged 65 and above) also differ in suicide risk when compared to depression in younger adults, with increased prevalence and risk for suicide associated with LLD^17^. Given that pro-inflammatory states also correlate with suicidal symptom presentation in MDD, suicidal ideation should also be considered in the context of depression and aging^18^.

Sex differences in MDD prevalence and associations with inflammatory immune peripheral markers suggest biological differences by sex in the manifestation of the disorder^19^. Periods of major hormonal changes during the lifespan of females - such as childbirth and menopause - may contribute to this differentiation^20^. Use of hormonal contraceptives and hormone replacement therapy (HRT), commonly prescribed to prevent pregnancy and to aide in menopausal symptom relief respectively, further complicates the relationship between hormonal fluctuations across the female lifespan, biomarkers, and depression. Studies have shown increased risk for depression and suicide associated with hormonal contraceptive use - especially during adolescence and in oral format^21,22^. Evidence for HRT-related depression is mixed, with a recent study identifying depression as a potential adverse side-effect of HRT dependent on when the therapy was initiated^23,24^.

Here, we use two large independent cohorts of mid- to late-life individuals to comprehensively model non-linear and interactive effects of age, sex, and genetic risk for depression on circulating peripheral biomarkers. We hypothesize that genetic liability for depression will be associated with levels of blood-based biomarkers more strongly in late life and will show sex-specific patterns of association.

## Materials and Methods

### Study datasets

We analyzed data from 362,074 individuals from the UK Biobank (age range: 39-72, mean=56.8 years; 53.7% female) and 22,965 individuals from the Canadian Longitudinal Study on Aging (CLSA; age range: 45-86, mean=63.0 years; 50.3% female). Written informed consent was provided by all participants. The UK Biobank is a large-scale biomedical database of over half a million volunteer participants from the United Kingdom^25^. The UK Biobank has approval from the North West Multi-centre Research Ethics Committee (https://www.hra.nhs.uk/about-us/committees-and-services/res-and-recs/search-research-ethics-committees/north-west-haydock/) and this study is approved by the UK Biobank under application ID 61530. CLSA is a longitudinal, nation-wide study of mid-through late-life Canadians^26^. CLSA has approval from 13 local research ethics boards across all participating sites (https://www.clsa-elcv.ca/governance/) and a data access agreement for this study is approved by CLSA under application number 2006026.

Details of genotyping and imputation for the UK Biobank and CLSA participants have been previously published^25,27^. Briefly, for the UK Biobank, genotyping was performed using the Applied Biosystems UK Biobank Axiom Array and imputed using the Haplotype Reference Consortium and UK10K + 1000 Genomes Phase 3 reference panels^28–30^. For CLSA, genotyping was performed using the Axiom 2.0 Assay Automated Workflow on Affymetrix NIMBUS protocol^27^. Samples were hybridized to UK Biobank arrays and imputed with TOPMed reference panels^31–33^. Both datasets were restricted to unrelated individuals of European genetic ancestry based on principal component analyses of genotypes (UK Biobank: https://biobank.ndph.ox.ac.uk/ukb/dset.cgi?id=2442; CLSA: https://www.clsa-elcv.ca/wp-content/uploads/2023/06/clsa_gwas_v3.pdf).

Peripheral biomarkers were measured at scale from blood samples collected at recruitment alongside demographic, lifestyle, and clinical information including medication burden, body-mass index (BMI), lifetime smoking status at time of recruitment (never, former, or current), and diagnoses for anxiety, depression, stroke, cancer, and Alzheimer’s disease. Diagnostic status in the UK Biobank was determined from the presence of ICD-10 codes retrieved from linked primary care or hospital inpatient data. Participants may also self-report a diagnosis at their baseline interview. For CLSA, diagnostic status as well as other covariates such as medication use and smoking status are self-reported either during the participant’s in-home, face-to-face interview or during their data collection site visit interview. More information on the variables included in this study can be found in **Supplementary Table 1** with descriptive statistics reported in **Supplementary Tables 2 and 3**.

### Peripheral biomarkers

We investigated 25 peripheral biomarkers available in both datasets that were measured from participants’ blood drawn at their initial assessment visit. Thirteen measures were derived from a complete blood count, including erythrocyte count and related measures (hemoglobin concentration, hematocrit percentage, mean corpuscular volume, mean corpuscular hemoglobin count and concentration, erythrocyte distribution width), mean thrombocyte volume and count, and leukocyte count and related measures (lymphocyte, monocyte, and granulocyte counts). Further differentiated counts of granulocytes (neutrophil, eosinophil, and basophil counts) were also available in the UK Biobank. The remaining ten measures broadly relate to metabolic function (albumin, alanine aminotransferase, and creatinine), lipid accumulation (cholesterol, high- and low-density lipoprotein, and triglyceride), overall health (glycated hemoglobin, vitamin D), and inflammatory signals (C-reactive protein; CRP). Individuals with levels beyond clinically acceptable ranges, following sex-specific adult (i.e., >18 years) guidelines from the Mayo Clinic (https://www.mayoclinic.org/**, Supplementary Table 4**), were excluded from analyses of that specific measure. Box-Cox transformation and normalization were applied to each measure using the *bestNormalize* (v1.9.0) package in R. Histograms of the measures before and after normalization can be found in **Supplementary Figures 1-4** and Box-Cox lambdas in **Supplementary Table 5**.

### Polygenic risk scoring

To calculate our polygenic risk scores for depression (PRS_dep_) in both datasets, we leveraged summary statistics from a genome-wide meta-analysis of both clinical and broadly-defined depression^13^. Since this meta-analysis included data from the UK Biobank, we used only the PGC29 summary statistics of major depressive disorder, which exclude UK Biobank samples, for PRS_dep_ calculations in the UK Biobank. For sensitivity analyses, we also calculated PGC29-only PRS_dep_ in CLSA. We applied the continuous shrinkage method (PRS-CS-auto: https://github.com/getian107/PRScs) to calculate the scores, which uses a Bayesian approach to define effect weights rather than discrete SNP selection. Specifically, we use the auto version that allows for the global shrinkage parameter, φ, to be learnt from the data, according to best practices for GWAS of highly polygenic traits with sufficient sample sizes as outlined by the authors of the package^34^. This Bayesian approach minimizes the number of parameters, requiring only the GWAS sample size, and thus does not require an independent test dataset for parameter tuning^34^. The 1000 Genomes Phase 3 European reference panel was used to account for linkage disequilibrium patterns in the data^30^. The top 10 genomic principal components were regressed out of the scores to account for fine population structure and standardized residuals were carried forward as the PRS_dep_ in all subsequent analyses.

### Statistical analysis

Analyses proceeded in four phases: 1) biomarker prioritization, 2) non-linear and interaction modelling, 3) sex-specific modelling, and 4) mediation analyses.

#### Biomarker prioritization

We tested for main effects of PRS_dep_ on measures of 25 peripheral biomarkers common to both the UK Biobank and CLSA. Linear regression models were fit to each outcome biomarker including main effects for PRS_dep_, sex, and non-linear age defined using cubic splines with three internal knots; interaction terms between age and sex; and selected covariates. Linear models fit with the UK Biobank data also include data collection site and fasting time prior to blood draw as covariates. Bonferroni correction for multiple testing was applied to each cohort separately for 25 tests in the UK Biobank (significant *p*<0.0020) and 23 in CLSA (significant *p*<0.0022). Biomarkers with observed significant main effects in both study cohorts were carried forward for the remaining statistical analyses.

#### Non-linear and interaction modelling

We incrementally tested interaction terms between non-linear age, sex, and PRS_dep_ with likelihood ratio tests to determine the effects of age and sex on the genetic effect. First, we compared non-linear age to linear age in simplified models without any interaction terms. Next, we investigated three interaction terms separately in addition to models with non-linear age: PRS_dep_-by-age, PRS_dep_-by-sex, and age-by-sex terms. Then, we fit models with two interaction terms: the age-by-sex term and additionally the PRS_dep_ interactions separately. Finally, we tested saturated models with all three interaction terms as well as the inclusion of a three-way interaction term. A total of nine nested models were investigated per biomarker.

#### Sex-specific exploration

We fit sex-stratified models including female-specific covariates to further investigate sex-specific effects. We first stratified the sample by sex and fit models separately in both the male and female subsamples, including self-reported menopause status, hormonal contraceptive use (specifically as an oral pill in the UK Biobank), hormone-replacement therapy status, and number of lifetime live births in the female subsample. We then further stratified the female subsample by menopause status and fit models for the pre-menopausal and post-menopausal subsamples, including age at which menopause was experienced in the post-menopausal subsample. Finally, *p*-values for each subgroup analyses are pooled across the UK Biobank and CLSA using Stouffer’s method.

#### Mediation analysis

We fit mediation models using the *mediation* (v4.5.0) package in R with PRS_dep_ as the independent variable to test bi-directional mediation between the biomarkers and depression diagnosis in both the UK Biobank and CLSA. We also investigate mediation through suicide-related thoughts and behaviors in the UK Biobank. Namely, we assessed mediation through binary responses to the following three questions: 1) “Ever thought that life was not worth living?”, 2) “Ever contemplated self-harm?”, and 3) “Ever self-harmed?”. Nonparametric bootstrapped confidence intervals were calculated with 1000 iterations of the mediation modelling.

## Results

### Biomarker prioritization

In the UK Biobank, elevated levels of five biomarkers (WBC, monocyte, neutrophil, lymphocyte, CRP) and decreased levels of two (creatinine and vitamin D) were found to be significantly associated with PRS_dep_ after accounting for non-linear age, sex, and their interaction; controlling for demographic, lifestyle, and clinical information; and correcting for 25 independent tests (i.e., 25 different biomarkers, *p*<0.0020; **Figure 1A**). Seven more biomarkers (triglyceride, albumin, basophil, RBC distribution width, MCH concentration, and alanine aminotransferase) were significantly associated with PRS_dep_ but did not survive multiple testing correction. All were elevated except RBC distribution width. In CLSA, elevated levels of six biomarkers (WBC, CRP, lymphocyte, granulocyte, glycated hemoglobin, and triglyceride) were significantly associated with PRS_dep_ with only triglyceride levels not surviving multiple testing correction (**Figure 1B**). Of these, the following five biomarkers showed consistent association with PRS_dep_ across both the UK Biobank and CLSA: WBC, neutrophil/granulocyte, lymphocyte, CRP, and triglycerides. Main effects of the PRS_dep_ and corresponding *p*-values and bootstrapped model *R*^2^ for each of the five outcomes and depression diagnosis are presented in **Table 1**.

**Figure 1:**
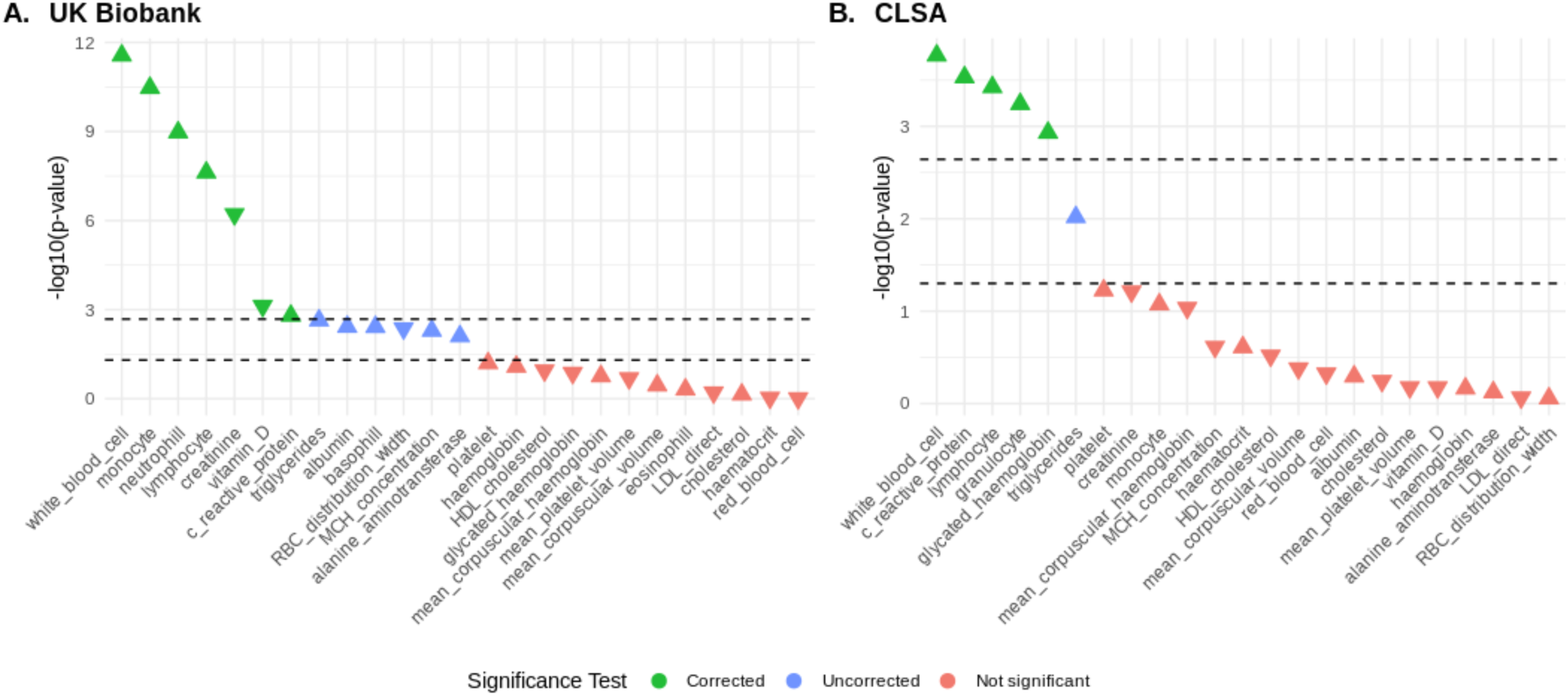
Points indicate the -log_10_(*p*-value) of main effects of PRS_dep_ in regression models in **A.** UK Biobank and **B.** CLSA with non-linear age and an age-by-sex interaction term. Shapes indicate direction of the effect and colors indicate significance levels. Bonferroni correction for multiple testing alongside uncorrected *p*-value < 0.05 thresholds shown by the dashed lines.

**Table 1:** Variance explained by regression models and main genetic effect for each outcome.

| Cohort | Outcome | Model Bootstrap $R^2$ | PRS<sub>dep</sub> Beta | PRS<sub>dep</sub> $p$-value |
| --- | --- | --- | --- | --- |
| UK Biobank | Depression | 0.17 | 0.14 | 1.10e-138 |
|  | White Blood Cell | 0.07 | 0.01 | 2.25e-12 |
|  | Neutrophil | 0.06 | 0.01 | 8.32e-10 |
|  | Lymphocyte | 0.06 | 9.60e-3 | 2.15e-8 |
|  | C-reactive Protein | 0.20 | 5.51e-3 | 7.50e-4 |
|  | Triglyceride | 0.10 | 6.72e-3 | 1.41e-3 |
| CLSA | Depression | 0.25 | 0.18 | 1.84e-18 |
|  | White Blood Cell | 0.10 | 0.03 | 1.71e-4 |
|  | Granulocyte | 0.10 | 0.02 | 5.73e-4 |
|  | Lymphocyte | 0.08 | 0.02 | 3.76e-4 |
|  | C-reactive Protein | 0.18 | 0.02 | 2.94e-4 |
|  | Triglyceride | 0.06 | 0.02 | 9.58e-3 |

**Model tested:** outcome ~ PRS + rcs(age,4) + sex + rcs(age, 4):sex + covariates
**Covariates:** medication burden + BMI + smoking status + anxiety + stroke + cancer + Alzheimer's disease
Covariates also include fasting time for analyses in the UK Biobank and depression for models where depression is not the outcome.
$R^2$ measures are corrected using the .632 bootstrap method with 100 repetitions.

### Non-linear and interaction modelling

In the UK Biobank, modeling age non-linearly provided better fitting models compared to a linear approach for all five biomarkers (32.80<χ^2^<1055.38; **Supplementary Table 6**). In CLSA, this was the case only for models of WBC count, granulocyte count, and triglyceride levels (26.69<χ^2^<158.11; **Supplementary Table 7**).

Regarding potential PRS_dep_ interaction effects, no consistent improvement in model fit is observed with the inclusion of either a PRS_dep_-by-age or PRS_dep_-by-sex interaction term. Furthermore, main effects of PRS_dep_ are largely diminished with the inclusion of any PRS_dep_ interactions (**Figure 2D-I**). Conversely, the inclusion of an age-by-sex interaction term provided the largest improvement across all models (22.40<χ^2^<2492.40; **Supplementary Tables 6**-**7**) except for models of CRP in CLSA (χ^2^=4.29, *p*=0.23). Main effects of PRS_dep_ also remained consistent with the inclusion of an age-by-sex interaction term (**Figure 2C**). Overall, models were significantly improved when age was modelled non-linearly and interacted with sex, and no meaningful interactions with PRS_dep_ were observed.

**Figure 2:**
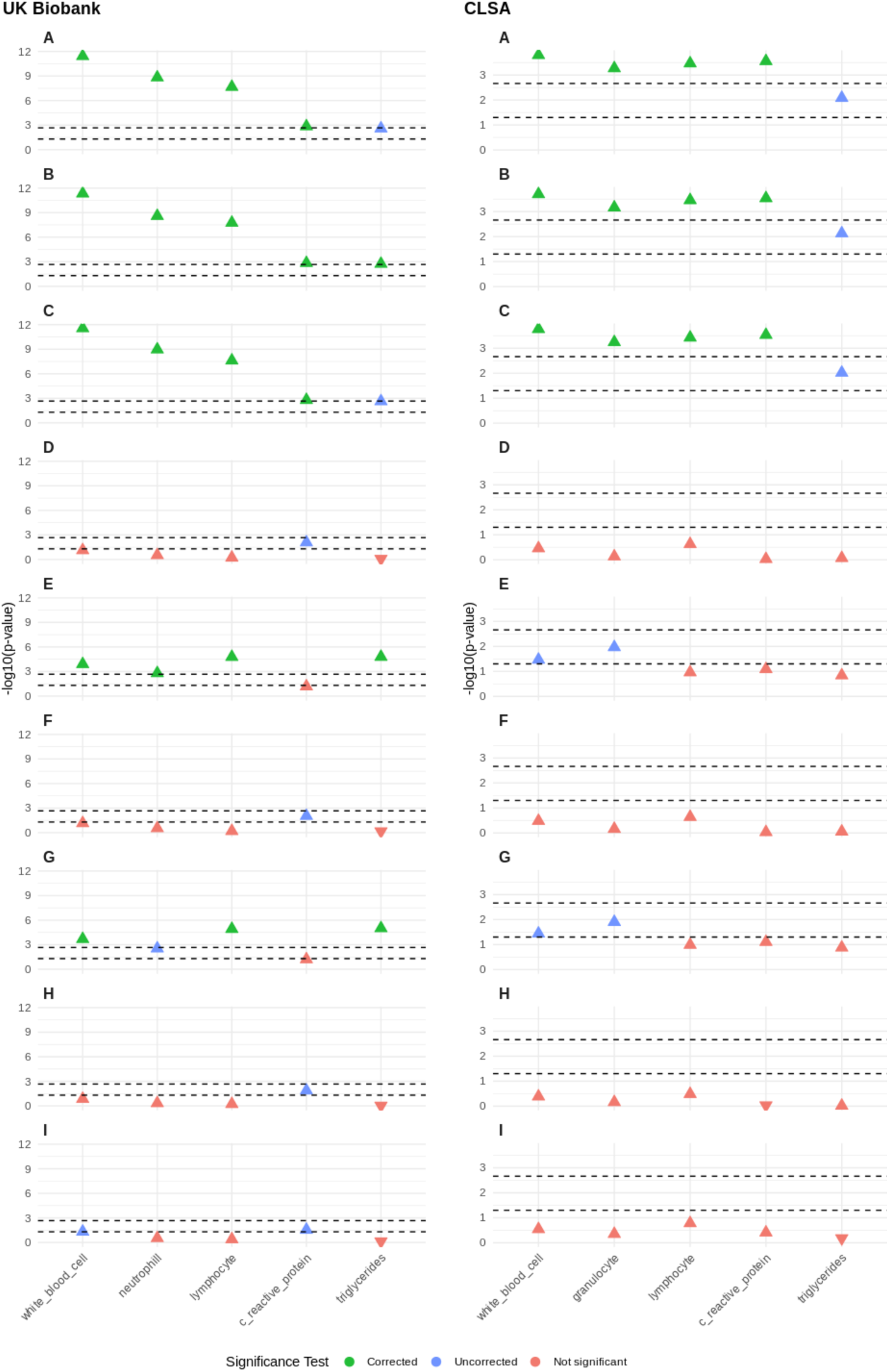
-log_10_(*p*-value) of main effects of PRS_dep_ in regression models with A) linear age, B) non-linear age, C) non-linear age and an age-by-sex interaction term, D) non-linear age and a PRS-by-age interaction term, E) non-linear age and a PRS-by-sex interaction term, F) non-linear age, age-by-sex, and PRS-by-age interaction terms, G) non-linear age, age-by-sex, and PRS-by-sex interaction terms, H) non-linear age, age-by-sex, PRS-by-age, and PRS-by-sex interaction terms, I) non-linear age, age-by-sex, PRS-by-age, and PRS-by-sex interaction terms and a three-way age-sex-PRS interaction term. Bonferroni correction for multiple testing alongside uncorrected *p*-value < 0.05 thresholds shown by the dashed lines.

To visualize these effects, we plotted model fits for each biomarker against age split by PRS_dep_ quintiles (**Figure 3**) and by sex (**Figure 4**). The main effects of PRS_dep_ (2.7×10^-^_12_<*p*<0.0096) and sex (3.7×10^-29^<*p*<0.0034) are significant across all biomarkers tested. There was no evidence for a PRS_dep_-by-age interaction; individuals in the highest quintile of genetic risk had consistently elevated levels of the biomarkers modeled when compared to individuals in lower quintiles of genetic risk. Furthermore, the non-linearity, when present, is driven primarily by the female subsample. Significant age-by-sex interaction effects were observed across all models except models of CRP levels and lymphocyte count in CLSA (**Table 2**).

**Figure 3:**
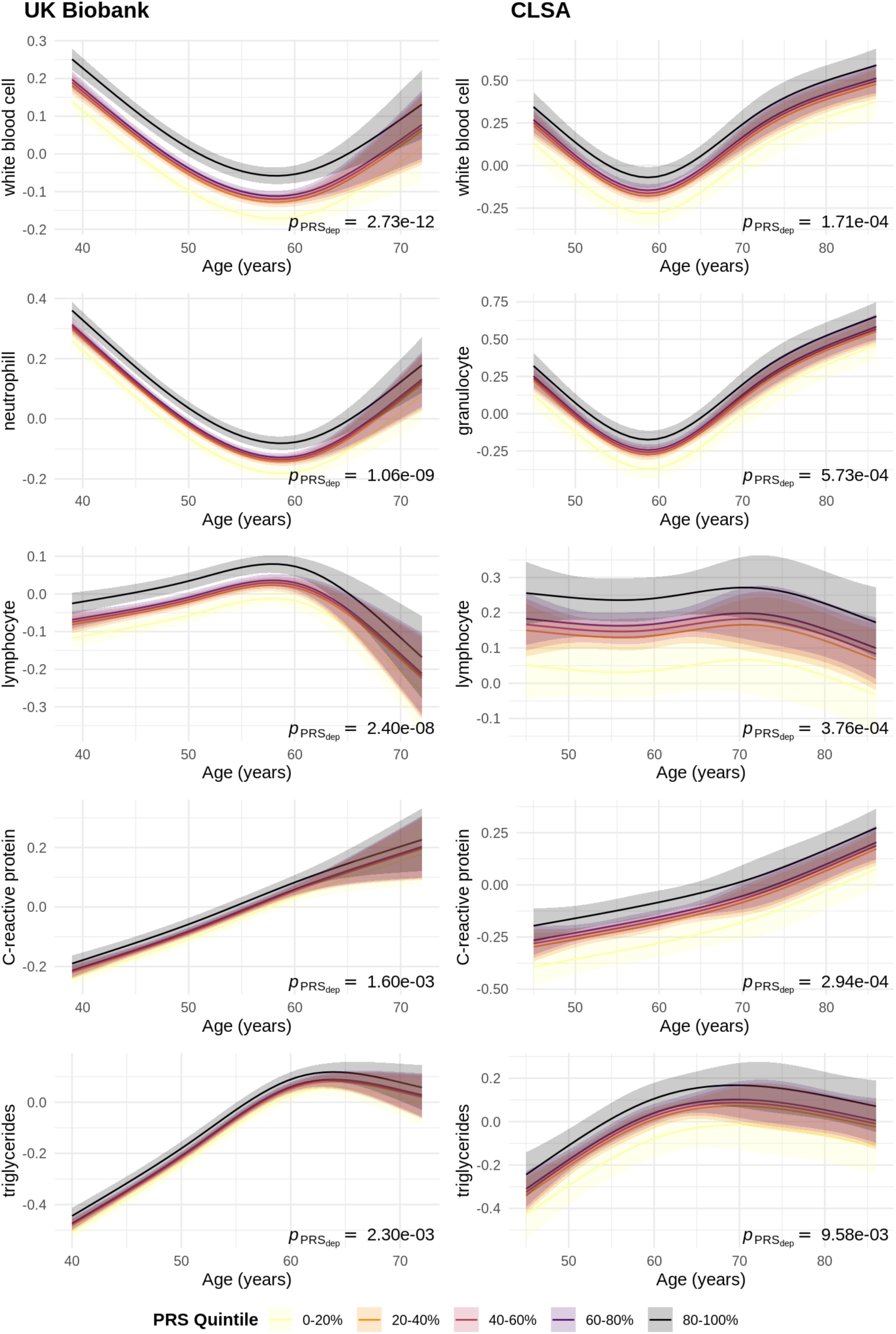
Interaction of age and PRS_dep_ on various blood biomarkers. Curves represent model fits (with margins for standard error) for the relationship between age and the biomarker for different quintiles of PRS_dep_ in the UK Biobank (left column) and CLSA (right column). Linear regression was used for modeling and all *p*-values are two-sided.

**Figure 4:**
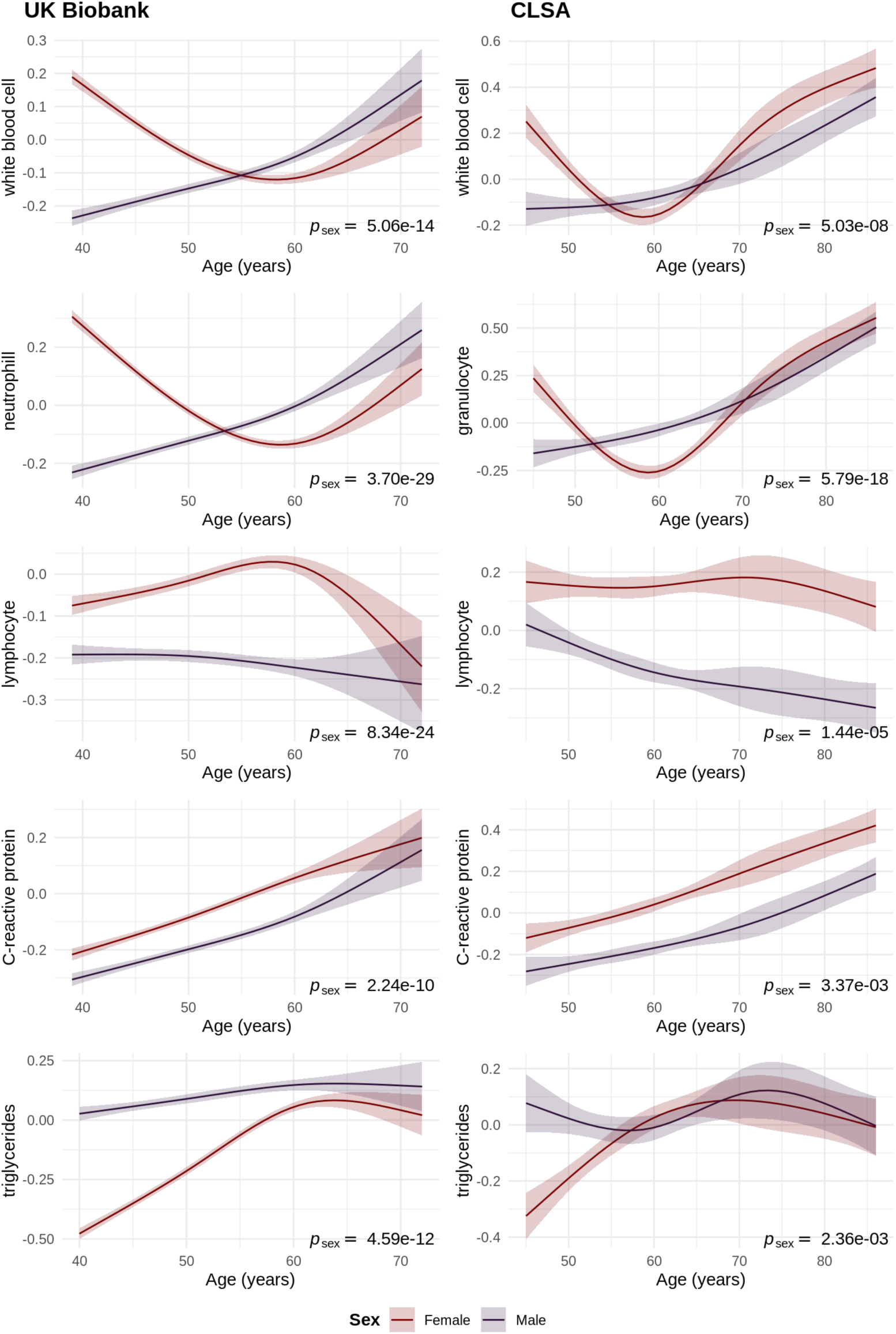
Interaction of age and sex on various blood biomarkers. Curves represent model fits (with margins for standard error) for the relationship between age and the biomarker for males and females separately in the UK Biobank (left column) and CLSA (right column). Linear regression was used for modeling and all *p*-values are two-sided.

**Table 2:** Non-linear age by sex effects for each outcome.

| Cohort | Outcome | Age<sup>1</sup>-sex<br>Beta | Age<sup>1</sup>-sex<br>p-value | Age<sup>2</sup>-sex<br>Beta | Age<sup>2</sup>-sex<br>p-value | Age<sup>3</sup>-sex<br>Beta | Age<sup>3</sup>-sex<br>p-value |
| --- | --- | --- | --- | --- | --- | --- | --- |
| UK<br>Biobank | Depression* | 0.06 | 0.20 | -0.04 | 0.71 | -0.15 | 0.78 |
|  | White Blood<br>Cell** | 0.25 | 1.59e-74 | -0.19 | 2.21e-09 | 0.21 | 0.16 |
|  | Neutrophil** | 0.33 | 1.08e-126 | -0.21 | 3.63e-11 | 0.09 | 0.55 |
|  | Lymphocyte*** | -0.04 | 8.87e-03 | -0.10 | 7.66e-04 | 0.63 | 4.77e-05 |
|  | C-reactive<br>Protein* | -0.01 | 0.34 | -0.05 | 0.11 | 0.29 | 0.05 |
|  | Triglyceride*** | -0.15 | 2.20e-16 | -0.10 | 0.02 | 0.63 | 3.58e-04 |
| CLSA | Depression* | 0.09 | 0.64 | -0.26 | 0.73 | 1.00 | 0.65 |
|  | White Blood<br>Cell*** | 0.44 | 2.42e-11 | -1.24 | 4.33e-07 | 3.15 | 8.27e-06 |
|  | Granulocyte*** | 0.58 | 9.12e-19 | -1.56 | 1.38e-10 | 3.90 | 2.77e-08 |
|  | Lymphocyte* | -0.10 | 0.13 | 2.74e-03 | 0.99 | 0.21 | 0.77 |
|  | C-reactive<br>Protein* | -0.03 | 0.67 | -0.07 | 0.77 | 0.30 | 0.66 |
|  | Triglyceride*** | -0.39 | 3.96e-06 | 0.87 | 5.16e-03 | -2.10 | 0.02 |

**Model tested:** outcome ~ PRS + rcs(age,4) + sex + rcs(age, 4):sex + covariates
**Covariates:** medication burden + BMI + smoking status + anxiety + stroke + cancer + Alzheimer's disease
Covariates also include fasting time for analyses in the UK Biobank and depression for models where depression is not the outcome.
\* no evidence for age-by-sex interactions
\*\* two of three non-linear age terms interact with sex
\*\*\* all three non-linear age terms interact with sex

### Sex-specific exploration

Higher genetic risk for depression was consistently associated with elevated WBC, neutrophil/granulocyte, lymphocyte, and CRP levels in males (**Table 3**). An association with elevated triglyceride levels was only observed in the male subgroup in CLSA (β=0.03; *p*=0.02) and not in the UK Biobank (*p*=0.92) and together this effect was not significant (Stouffer’s *p*=0.32).

**Table 3:** Variance explained by regression models and main genetic effect for each biomarker for different subsamples.

| Biomarker | Sub-sample | UK Biobank |  |  |  | CLSA |  |  |  | Stouffer's *p*-value |
| --- | --- | --- | --- | --- | --- | --- | --- | --- | --- | --- |
|  |  | Sample size | Variance explained | Standardized beta of PRS | *p*-value of PRS | Sample size | Variance explained | Standardized beta of PRS | *p*-value of PRS |  |
| white blood cell | male | 149400 | 0.082 | 0.014 | 7.17E-09*** | 9853 | 0.106 | 0.028 | 3.16e-03** | 1.43E-09*** |
|  | female | 145129 | 0.079 | 0.008 | 1.70E-03*** | 6675 | 0.102 | 0.018 | 1.24e-01 | 1.94E-03*** |
|  | pre-menopause | 38213 | 0.065 | 0.004 | 4.50E-01 | 1335 | 0.072 | 0.030 | 2.74e-01 | 3.04E-01 |
|  | post-menopause | 100089 | 0.082 | 0.009 | 1.72E-03*** | 5184 | 0.112 | 0.016 | 1.99e-01 | 3.83E-03** |
| neutrophil/granulocyte | male | 148920 | 0.065 | 0.013 | 1.11E-07*** | 9978 | 0.095 | 0.021 | 2.92e-02* | 2.85E-07*** |
|  | female | 145433 | 0.070 | 0.007 | 3.39E-03** | 6830 | 0.100 | 0.021 | 6.21e-02 | 1.34E-03*** |
|  | pre-menopause | 37855 | 0.048 | 0.001 | 8.39E-01 | 1373 | 0.074 | 0.060 | 2.82e-02* | 2.58E-01 |
|  | post-menopause | 100725 | 0.067 | 0.010 | 9.13E-04*** | 5299 | 0.104 | 0.013 | 3.01e-01 | 5.05E-03** |
| lymphocyte | male | 150730 | 0.056 | 0.008 | 1.43E-03*** | 9925 | 0.072 | 0.031 | 1.32e-03*** | 1.15E-05*** |
|  | female | 145268 | 0.050 | 0.010 | 1.33E-04*** | 6589 | 0.057 | 0.017 | 1.51e-01 | 4.68E-04*** |
|  | pre-menopause | 39728 | 0.058 | 0.014 | 4.04E-03** | 1344 | 0.077 | -0.030 | 2.40e-01 | 8.84E-03** |
|  | post-menopause | 98764 | 0.047 | 0.009 | 5.58E-03** | 5093 | 0.058 | 0.033 | 1.17e-02** | 3.40E-04*** |
| C-reactive protein | male | 142317 | 0.156 | 0.007 | 3.66E-03** | 10142 | 0.150 | 0.029 | 1.08e-03*** | 2.40E-05*** |
|  | female | 135050 | 0.237 | 0.003 | 2.36E-01 | 6602 | 0.225 | 0.011 | 3.11e-01 | 1.96E-01 |
|  | pre-menopause | 37165 | 0.236 | 0.005 | 2.95E-01 | 1324 | 0.280 | 0.017 | 4.92e-01 | 3.46E-01 |
|  | post-menopause | 91675 | 0.223 | 0.002 | 5.02E-01 | 5127 | 0.209 | 0.009 | 4.80e-01 | 4.87E-01 |
| triglycerides | male | 79218 | 0.052 | 0.000 | 9.15E-01 | 6089 | 0.049 | 0.029 | 2.15e-02* | 3.22E-01 |
|  | female | 106309 | 0.124 | 0.012 | 1.81E-05*** | 4851 | 0.093 | 0.005 | 6.94e-01 | 5.19E-03** |
|  | pre-menopause | 33448 | 0.105 | 0.014 | 8.79E-03** | 1101 | 0.113 | 0.043 | 1.36e-01 | 7.00E-03** |
|  | post-menopause | 68252 | 0.080 | 0.013 | 2.59E-04*** | 3638 | 0.071 | -0.003 | 8.29e-01 | 3.73E-02* |

\*\*\*= $p$ -value < 0.05 /20; \*\*= $p$ -value < 0.05/4; \*= $p$ -value < 0.05
Stouffer's $p$ -values were calculated across the two populations per subgrouping per biomarker.

Across the female subgroups, the effects of PRS_dep_ on WBC and neutrophil/granulocyte counts were only observed in females who reported already having experienced menopause (WBC: Stouffer’s *p*=3.83×10^-3^; neutrophil/granulocyte: Stouffer’s *p*=5.05×10^-3^). Elevated lymphocyte and triglyceride levels were associated with higher genetic risk for depression in females regardless of menopause status. No effects on CRP levels were observed for females across either cohort.

### Mediation analysis

Across all biomarkers tested in the UK Biobank, the effect of PRS_dep_ showed evidence for mediation through a depression diagnosis (**Table 4**). This mediation was found to be bi-directional wherein the association between PRS_dep_ and a depression diagnosis was also mediated by each of the biomarkers. In particular, the proportion mediated by a depression diagnosis in mediation models of lymphocyte count was negative (proportion mediated = - 0.05%), suggesting a small suppression effect. The largest mediation observed was through a depression diagnosis between PRS_dep_ and triglyceride levels (proportion mediated = 12.6%, CI_95%_=[7.6%, 30%]) followed by CRP levels (proportion mediated = 10.7%, CI_95%_=[7.2%, 22%]). Mediation through depression diagnosis in CLSA was only present in lymphocyte (proportion mediated = 6.4%, CI_95%_=[1.8%, 15%]) and CRP (proportion mediated = 4.6%, CI_95%_=[0.7%, 13%]) models (**Supplementary Table 8**).

**Table 4:** Mediation models testing bidirectional mediation through a depression diagnosis in the UK Biobank.

| Predictor | → | Mediator | → | Outcome | Mediated | Direct | Proportion | Total |
| --- | --- | --- | --- | --- | --- | --- | --- | --- |
| PRS | → | MDD | → | WBC | $3.17 \times 10^{-4}$ | 0.0140 | 0.0221 | 0.0143 |
| PRS | → | WBC | → | MDD | $2.98 \times 10^{-5}$ | 0.0187 | 0.0016 | 0.0187 |
| PRS | → | MDD | → | Neutrophil | $5.69 \times 10^{-4}$ | 0.0127 | 0.0428 | 0.0133 |
| PRS | → | Neutrophil | → | MDD | $4.48 \times 10^{-5}$ | 0.0186 | 0.0024 | 0.0186 |
| PRS | → | MDD | → | Lymphocyte | $-5.37 \times 10^{-4}$ | 0.0093 | -0.0613 | 0.0088 |
| PRS | → | Lymphocyte | → | MDD | $-2.46 \times 10^{-5}$ | 0.0189 | -0.0013 | 0.0189 |
| PRS | → | MDD | → | CRP | $7.42 \times 10^{-4}$ | 0.0062 | 0.1065 | 0.0070 |
| PRS | → | CRP | → | MDD | $4.14 \times 10^{-5}$ | 0.0181 | 0.0023 | 0.0181 |
| PRS | → | MDD | → | Triglyceride | $9.36 \times 10^{-4}$ | 0.0065 | 0.1259 | 0.0074 |
| PRS | → | Triglyceride | → | MDD | $4.62 \times 10^{-5}$ | 0.0178 | 0.0026 | 0.0179 |

Evidence of mediation by self-harm behaviors was observed only for responses to the most general question “Ever thought that life was not worth living?” in models of triglyceride levels (proportion mediated = 6.2%, CI_95%_=[2.5%, 17%]), neutrophil count (proportion mediated = 4.5%, CI_95%_=[0.98%, 10%]), and WBC count (proportion mediated = 2.8%, CI_95%_=[0.34%, 6%]) (**Supplementary Table 9**). No mediation was observed for responses to more granular self-harm questions tested.

## Discussion

Our results primarily highlight robust associations between genetic risk for depression and inflammatory peripheral biomarkers in mid-late life. After assessing 25 biomarkers and controlling for multiple tests, PRSs for depression remain associated with elevated levels of white blood cell, granulocyte (specifically neutrophil), and lymphocyte counts, and C-reactive protein and triglyceride levels despite modelling non-linearities in age to account for variation throughout the human lifespan, as well as interactions with sex to account for sex-specific experiences such as menopause. These results were concordant across two large independent cohorts (UK Biobank: n=337,321; CLSA: n=22,965), indicating that these findings are not study specific.

WBC, granulocyte, lymphocyte, and CRP are all indices of inflammation and have previously been independently associated with major depressive disorder and depressive symptoms^35,36^. Elevated triglyceride levels have also been implicated in activation of inflammatory mediators, and accumulation as a response to pathogen-activated inflammation^37,38^. These findings suggest that a genetic predisposition for depression may also confer vulnerability for inflammation. Previous studies have investigated this relationship in the other direction, identifying contributions of inflammation-related genetics to depression (i.e., studying genetic variants in immune genes and mRNA expression)^10,39^. In this study, we show consistent association between genetic risk for depression and inflammatory biomarkers across the lifespan, but specific to subgroups of individuals.

Sex-differences in depression is a well-known phenomenon with prevalence rates estimated to be twice as high in females. Despite this, little is known about the biological mechanisms that underly the sex-specific etiology of depression^40^. Recent work have considered immunology and inflammation as potential causes of this discrepancy^41,42^. For example, Shafiee et al. observed associations between depression and elevated levels of WBC in males and not in females^43^. This contrasts with our findings where WBC count was elevated regardless of sex. However, we found that this association was only present in post-menopausal females, consistent with studies showing elevated inflammatory markers as a result of menopause^44–46^. Shafiee et al. did not include any information on menopausal status in their study and investigated a much younger cohort (aged 35-65 years). Furthermore, we identified male-specific effects of CRP across both investigated cohorts, suggesting that elevated levels of CRP as a response to depression may be modulated differently in females. Population-based studies of CRP in depressed individuals report inconsistent sex-specificity that is likely tied to depression severity. In both clinically obese and general urban-dwelling populations, elevated levels of CRP has been associated with depressive symptoms^47,48^. However, in adults with moderate to severe major depressive disorder, elevated CRP is associated with symptom severity only in females, further reinforcing the heterogeneity of this condition^49^.

We further examined the effects of a depression diagnosis and suicide-related thoughts and behaviours through mediation models and found that the association between PRS_dep_ and CRP is mediated by a depression diagnosis, but not by a lack of will to live. This suggests that the mediated effect through CRP may be specific to the behavioural components of a depression diagnosis, such as lethargy and fatigue, rather than psychiatric components such as anhedonia and suicidality^50,51^. Triglyceride levels, however, are mediated by both depression diagnosis and suicidality, suggesting that elevated triglyceride levels can indicate more than a manifestation of sedentary behavior and depression^8,52^. All our mediation models suggest that mediations by the biomarkers are much weaker than mediations by a depression diagnosis.

We acknowledge that the source summary statistics used to develop our polygenic risk scores differ between the UK Biobank and CLSA. This was done to remove bias that occurs in the SNP selection when the source GWAS includes the population under investigation (as is the case for the UK Biobank in this study). We chose to retain the UK Biobank summary statistics for our CLSA PRS to utilize the largest and most up-to-date summary statistics available for depression. Our sensitivity analysis for UK Biobank-exclusive PRS in CLSA show no significant changes in results. Furthermore, our criteria for clinically acceptable ranges for the biomarkers tested in this study are arbitrary. We chose to rely mostly on the Mayo Clinic’s guidelines, but these thresholds may vary across clinical institutions and circumstances.

Overall, we identified robust associations between genetic risk for depression and peripheral biomarkers consistent with inflammation. We showed that these associations are present throughout the lifespan but manifest in a sex-specific manner. Stratification by more granular sex-specific factors, such as menopause status, showed different patterns of associations between inflammatory markers and genetic risk for depression. Biomarker specificity to subgroups of individuals can guide targeted treatment and intervention. Our mediation analyses suggest that up to 13% (e.g., triglyceride levels) of the associations observed may be accounted for by a depression diagnosis, but not the other way around. Considering this potential sex-dependency, these findings should encourage efforts to uncover genome-wide markers of sex-specific depression, which are needed to fully understand these sex-related interaction effects.

## Data Availability

All data produced in the present work are contained in the manuscript.

## Additional Information

This research has been conducted using the UK Biobank Resource under Application Number 61530. This research was made possible using the data/biospecimens collected by the Canadian Longitudinal Study on Aging (CLSA). Funding for the Canadian Longitudinal Study on Aging (CLSA) is provided by the Government of Canada through the Canadian Institutes of Health Research (CIHR) under grant reference: LSA 94473 and the Canada Foundation for Innovation, as well as the following provinces, Newfoundland, Nova Scotia, Quebec, Ontario, Manitoba, Alberta, and British Columbia. This research has been conducted using the CLSA dataset [Genome-wide Genetic Data Release version 3.0; Comprehensive Baseline Dataset Version 7.0], under Application Number [2006026].

