## Supplementary for "Investigation of sex- and age-specific effects of genetic risk for depression on peripheral biomarkers"

### Tables

**Table 1:** UK Biobank field IDs and CLSA variable names of the variables included in this study.

| Measure | UK Biobank Field ID | CLSA Variable Name |
|---|---|---|
| White blood cell (leukocyte) count | 30000 | BLD_WBC_COM |
| Lymphocyte count | 30120 | BLD_LY_NB_COM |
| Monocyte count | 30130 | BLD_MO_NB_COM |
| Neutrophil count | 30140 | - |
| Eosinophil count | 30150 | - |
| Basophil count | 30160 | - |
| Granulocyte count | - | BLD_GR_NB_COM |
| Red blood cell (erythrocyte) count | 30010 | BLD_RBC_COM |
| Hemoglobin concentration | 30020 | BLD_Hgb_COM |
| Hematocrit percentage | 30030 | BLD_Hct_COM |
| Mean corpuscular volume | 30040 | BLD_MCV_COM |
| Mean corpuscular hemoglobin | 30050 | BLD_MCH_COM |
| Mean corpuscular hemoglobin concentration | 30060 | BLD_MCHC_COM |
| Red blood cell (erythrocyte) distribution width | 30070 | BLD_RDW_COM |
| Platelet (thrombocyte) count | 30080 | BLD_Plt_COM |
| Mean platelet (thrombocyte) volume | 30100 | BLD_MPV_COM |
| Albumin | 30600 | BLD_ALB_COM |
| Alanine aminotransferase | 30620 | BLD_ALT_COM |
| Creatinine | 30700 | BLD_CREAT_COM |
| Cholesterol | 30690 | BLD_CHOL_COM |
| HDL cholesterol | 30760 | BLD_HDL_COM |
| LDL direct | 30780 | BLD_LDL_COM |
| Triglyceride | 30870 | BLD_TRIG_COM |
| Glycated hemoglobin (HbA1c) | 30750 | BLD_HBA1c_COM |
| Vitamin D | 30890 | BLD_VITD_COM |
| C-reactive protein | 30710 | BLD_HSCRP_COM |
| Sex | 31 | SEX_ASK_COM |
| Age | 21022 | AGE_NMBR_COM |
| Depression | 130895 | DPR_CLINDEP_COM |
| Anxiety | 130907 | CCC_ANXI_COM |
| Stroke | 42007 | CCC_CVA_COM |
| Cancer | 2453 | CCC_CANC_COM |
| Alzheimer's disease | 131037 | CCC_ALZH_COM |
| Smoking status | 20116 | SMK_DSTY_COM |
| Medication burden* | 137 | OAR_MED_COM, HYP_UTHYRMED_COM, STR_MED_COM, HBP_MED_COM, DIA_MED_COM, IHD_MED_COM, HYP_OTHYRMED_COM, STR_TIAMED_COM, DPR_MED_COM, OST_MED_COM, PKD_MED_COM, CAO_MED_COM, ICQ_EMBMED_COM |
| Body mass index | 21001 | HWT_DBMI_COM |
| Menopause status | 2724 | WHO_MENOP_COM |

| | | |
|---|---|---|
| Age at menopause | 3581 | WHO_MPAG_AG_COM |
| Number of live births | 2734 | WHO_PREG_LIVE_NB_COF1 |
| Ever taken oral contraceptive pill | 2784 | - |
| Ever used hormonal contraceptives | - | WHO_CONCP_COF1 |
| Ever used hormone-replacement therapy | 2814 | WHO_HRT_COM |
| Fasting time | 74 | - |
| Assessment centre | 54 | - |

*Medication burden in the CLSA was calculated as a count of "yes" responses across the listed variables when asked if the participant was currently taking medication for specific diseases.
More information on all listed variables can be found through the respective data showcase websites (UK Biobank: https://biobank.ndph.ox.ac.uk/showcase/index.cgi; CLSA: https://datapreview.clsa-elcv.ca/datasets)

**Table 2:** Descriptive statistics of peripheral biomarkers demographics of 337,321 individuals from the UK Biobank included in the study.

| Measure | Male (*n*=167,569) | | Female (*n*=194,505) | | Test stat | *p*-value |
|---|---|---|---|---|---|---|
| | Range/count | Mean (SD)/% | Range/count | Mean (SD)/% | | |
| White blood cell (leukocyte) count ($10^9$/L) | 3.40-9.60 | 6.61 (1.35) | 3.40-9.60 | 6.58 (1.35) | 6.8 | 7.40e-12 |
| Lymphocyte count ($10^9$/L) | 0.95-3.07 | 1.83 (0.47) | 0.95-3.07 | 1.92 (0.48) | -54.7 | < 2.2e-16 |
| Monocyte count ($10^9$/L) | 0.26-0.81 | 0.50 (0.13) | 0.26-0.81 | 0.45 (0.12) | 118.9 | < 2.2e-16 |
| Neutrophil count ($10^9$/L) | 1.56-6.45 | 4.04 (1.04) | 1.56-6.45 | 4.00 (1.04) | 13.1 | 3.23e-39 |
| Eosinophil count ($10^9$/L) | 0.03-0.48 | 0.17 (0.10) | 0.03-0.48 | 0.16 (0.09) | 56.5 | < 2.2e-16 |
| Basophil count ($10^9$/L) | 0.01-0.08 | 0.03 (0.02) | 0.01-0.08 | 0.03 (0.02) | -8.5 | 2.87e-17 |
| Red blood cell (erythrocyte) count ($10^{12}$/L) | 4.35-5.65 | 4.82 (0.28) | 3.92-5.13 | 4.38 (0.26) | 452.6 | < 2.2e-16 |
| Hemoglobin concentration (g/dL) | 13.20-16.60 | 15.00 (0.78) | 11.60-15.00 | 13.49 (0.75) | 558.0 | < 2.2e-16 |
| Hematocrit percentage (%) | 38.30-48.60 | 43.40 (2.34) | 35.50-44.90 | 39.61 (2.14) | 477.0 | < 2.2e-16 |
| Mean corpuscular volume (fL) | 78.20-97.90 | 91.12 (3.46) | 78.20-97.93 | 90.79 (3.62) | 26.2 | 1.39e-150 |
| Mean corpuscular hemoglobin (pg/cell) | 27.00-31.00 | 30.10 (0.78) | 27.00-31.00 | 29.97 (0.86) | 28.4 | 1.08e-176 |
| Mean corpuscular hemoglobin concentration (g/dL) | 32.00-36.00 | 34.51 (0.76) | 32.00-36.00 | 34.34 (0.78) | 61.8 | < 2.2e-16 |
| Red blood cell (erythrocyte) distribution width (%) | 11.80-14.50 | 13.28 (0.55) | 12.20-16.10 | 13.44 (0.72) | -73.3 | < 2.2e-16 |
| Platelet (thrombocyte) count ($10^9$/L) | 135.00-317.00 | 230.68 (40.65) | 157.00-371.00 | 261.31 (47.15) | -198.7 | < 2.2e-16 |
| Mean platelet (thrombocyte) volume (fL) | 7.00-9.00 | 8.38 (0.45) | 7.00-9.00 | 8.40 (0.44) | -8.0 | 1.77e-15 |
| Albumin (g/dL) | 35.00-50.00 | 45.28 (2.33) | 35.06-50.00 | 44.82 (2.38) | 54.1 | < 2.2e-16 |
| Alanine aminotransferase (U/L) | 7.00-55.00 | 25.19 (9.45) | 7.00-45.00 | 18.88 (7.07) | 214.0 | < 2.2e-16 |
| Creatinine (μmol/L) | 65.40-119.30 | 82.39 (10.48) | 52.20-91.90 | 65.33 (8.26) | 496.2 | < 2.2e-16 |
| Cholesterol (mmol/L) | 0.60-5.20 | 4.43 (0.58) | 1.88-5.20 | 4.60 (0.48) | -54.4 | < 2.2e-16 |
| HDL cholesterol (mmol/L) | 1.00-4.19 | 1.36 (0.28) | 1.30-4.40 | 1.73 (0.32) | -308.3 | < 2.2e-16 |
| LDL direct (mmol/L) | 0.27-3.40 | 2.76 (0.46) | 0.75-3.40 | 2.84 (0.41) | -37.1 | 7.08e-300 |
| Triglyceride (mmol/L) | 0.23-1.70 | 1.17 (0.31) | 0.23-1.70 | 1.11 (0.31) | 48.6 | < 2.2e-16 |
| Glycated hemoglobin (HbA1c) (mmol/mol) | 20.00-48.00 | 35.12 (3.92) | 20.00-48.00 | 35.06 (3.74) | 4.9 | 1.09e-06 |
| Vitamin D (ng/mL) | 10.00-80.00 | 45.87 (16.90) | 10.00-80.00 | 45.95 (17.03) | -1.4 | 1.74e-01 |
| C-reactive protein (mg/L) | 0.08-5.00 | 1.46 (1.10) | 0.08-5.00 | 1.51 (1.17) | -10.8 | 2.49e-27 |
| Age (years) | 39.00-72.00 | 57.06 (8.10) | 40.00-71.00 | 56.62 (7.92) | 16.5 | 3.94e-61 |
| Depression (yes) | 15382 | 9.18 | 27100 | 13.93 | 1963 | < 2.2e-16 |
| Depression (no) | 152187 | 90.82 | 167405 | 86.07 | | |
| Anxiety (yes) | 7853 | 4.69 | 14540 | 7.48 | 1206 | 2.63e-264 |
| Anxiety (no) | 159716 | 95.31 | 179965 | 92.52 | | |
| Stroke (yes) | 5573 | 3.33 | 3878 | 1.99 | 628 | 1.54e-138 |
| Stroke (no) | 161996 | 96.67 | 190627 | 98.01 | | |
| Cancer (yes) | 10482 | 6.27 | 17932 | 9.26 | 1104 | 5.18e-242 |
| Cancer (no) | 156680 | 93.73 | 175753 | 90.74 | | |
| Alzheimer's disease (yes) | 830 | 0.50 | 879 | 0.45 | 4 | 0.0607 |
| Alzheimer's disease (no) | 166739 | 99.50 | 193626 | 99.55 | | |
| Smoking status (current) | 20107 | 12.05 | 17027 | 8.79 | 3914 | < 2.2e-16 |
| Smoking status (former) | 65373 | 39.18 | 62311 | 32.16 | | |

| | | | | | | |
|---|---|---|---|---|---|---|
| Smoking status (never) | 81392 | 48.78 | 114402 | 59.05 | | |
| Medication burden (median) | 0-28 | 2 | 0-48 | 2 | 1304 | 1.31e-285 |
| Body mass index (kg/m$^2$) | 12.81-68.41 | 27.84 (4.25) | 12.12-74.68 | 27.00 (5.15) | 53.7 | < 2.2e-16 |
| Menopause (yes) | - | - | 119473 | 73.16 | - | - |
| Menopause (no) | - | - | 43834 | 26.84 | - | - |
| Age at menopause (years) | - | - | 18.00-68.00 | 49.74 (5.11) | - | - |
| Number of live births | - | - | 0-22 | 1.822 (1.20) | - | - |
| Ever taken oral contraceptive pill (yes) | - | - | 220301 | 81.05 | - | - |
| Ever taken oral contraceptive pill (no) | - | - | 51505 | 18.49 | - | - |
| Ever used hormone-replacement therapy (yes) | - | - | 103884 | 38.24 | - | - |
| Ever used hormone-replacement therapy (no) | - | - | 167765 | 61.75 | - | - |
| Fasting time (hours) | 0.00-72.00 | 3.86 (2.53) | 0.00-48.00 | 3.67 (2.22) | 23.2 | 4.51e-119 |

Welch's two sample t-test was used to compare means of continuous variables and Pearson's Chi-squared test with Yates' continuity correction for counts of categorical variables. Kruskal-Wallis rank sum test was used for medication burden specifically due to the non-normal distribution of count data. All *p*-values are two-sided.

**Table 3:** Descriptive statistics of peripheral biomarkers and demographics of 22,965 individuals from the CLSA included in the study.

| Measure | Male (*n*=11,410) | | Female (*n*=11,555) | | Test stat | *p*-value |
|---|---|---|---|---|---|---|
| | Range/count | Mean (SD)/% | Range/count | Mean (SD)/% | | |
| White blood cell (leukocyte) count ($10^9$/L) | 3.40-9.60 | 6.44 (1.36) | 3.40-9.60 | 6.42 (1.35) | 1.0 | 0.305 |
| Lymphocyte count ($10^9$/L) | 1.00-3.00 | 1.85 (0.46) | 1.00-3.00 | 1.97 (0.46) | -19.6 | 2.38e-84 |
| Monocyte count ($10^9$/L) | 0.30-0.80 | 0.40 (0.11) | 0.30-0.80 | 0.39 (0.11) | 8.5 | 2.52e-17 |
| Granulocyte count ($10^9$/L) | 1.60-7.00 | 4.25 (1.13) | 1.60-7.00 | 4.11 (1.12) | 8.7 | 3.75e-18 |
| Red blood cell (erythrocyte) count ($10^{12}$/L) | 4.35-5.65 | 4.89 (0.31) | 3.92-5.13 | 4.46 (0.29) | 96.9 | < 2.2e-16 |
| Hemoglobin concentration (g/dL) | 132.00-166.00 | 148.32 (8.35) | 116.00-150.00 | 133.98 (8.03) | 121.1 | < 2.2e-16 |
| Hematocrit percentage (%) | 0.38-0.49 | 0.44 (0.02) | 0.36-0.45 | 0.40 (0.02) | 102.3 | < 2.2e-16 |
| Mean corpuscular volume (fL) | 78.20-97.90 | 91.95 (3.41) | 78.20-97.90 | 91.48 (3.54) | 9.5 | 3.43e-21 |
| Mean corpuscular hemoglobin (pg/cell) | 27.00-31.00 | 29.74 (0.93) | 27.00-31.00 | 29.62 (0.98) | 7.5 | 5.10e-14 |
| Mean corpuscular hemoglobin concentration (g/dL) | 320.00-360.00 | 333.62 (7.97) | 320.00-360.00 | 332.46 (7.66) | 10.2 | 2.59e-24 |
| Red blood cell (erythrocyte) distribution width (%) | 11.80-14.50 | 13.17 (0.59) | 12.20-16.10 | 13.44 (0.80) | -26.9 | 9.02e-157 |
| Platelet (thrombocyte) count ($10^9$/L) | 135.00-317.00 | 207.22 (40.85) | 157.00-371.00 | 238.86 (47.05) | -50.6 | < 2.2e-16 |
| Mean platelet (thrombocyte) volume (fL) | 7.00-9.00 | 8.05 (0.54) | 7.00-9.00 | 8.08 (0.53) | -3.7 | 2.16e-04 |
| Albumin (g/dL) | 35.00-50.00 | 40.19 (2.46) | 35.00-50.00 | 39.83 (2.48) | 10.8 | 4.27e-27 |
| Alanine aminotransferase (U/L) | 7.00-55.00 | 24.45 (8.98) | 7.00-45.00 | 19.69 (6.93) | 44.0 | < 2.2e-16 |
| Creatinine (μmol/L) | 66.00-119.00 | 89.02 (11.60) | 53.00-91.00 | 70.34 (8.78) | 130.8 | < 2.2e-16 |
| Cholesterol (mmol/L) | 1.80-5.20 | 4.18 (0.67) | 2.01-5.20 | 4.47 (0.55) | -25.5 | 2.48e-139 |
| HDL cholesterol (mmol/L) | 1.00-4.15 | 1.43 (0.35) | 1.30-4.45 | 1.84 (0.42) | -71.0 | < 2.2e-16 |
| LDL direct (mmol/L) | 0.02-3.40 | 2.31 (0.66) | 0.08-3.40 | 2.49 (0.60) | -18.2 | 6.66e-73 |
| Triglyceride (mmol/L) | 0.24-1.70 | 1.16 (0.32) | 0.30-1.70 | 1.14 (0.31) | 2.8 | 4.60e-03 |
| Glycated hemoglobin (HbA1c) (%) | 4.30-6.50 | 5.50 (0.34) | 4.20-6.50 | 5.46 (0.33) | 8.8 | 1.11e-18 |
| Vitamin D (ng/mL) | 10.00-80.00 | 57.73 (15.77) | 10.00-80.00 | 58.48 (15.78) | -2.3 | 0.0212 |
| C-reactive protein (mg/L) | 0.10-5.00 | 1.42 (1.09) | 0.10-5.00 | 1.58 (1.19) | -9.8 | 9.09e-23 |
| Age (years) | 45.00-86.00 | 63.27 (10.17) | 45.00-86.00 | 62.79 (10.13) | 3.5 | 4.12e-04 |
| Depression (yes) | 1368 | 12.04 | 2410 | 20.94 | 328 | 2.38e-73 |
| Depression (no) | 9997 | 87.96 | 9098 | 79.06 | | |
| Anxiety (yes) | 745 | 6.55 | 1243 | 10.79 | 129 | 5.94e-30 |
| Anxiety (no) | 10629 | 93.45 | 10276 | 89.21 | | |
| Stroke (yes) | 234 | 2.06 | 143 | 1.24 | 23 | 1.57e-06 |
| Stroke (no) | 11136 | 97.94 | 11377 | 98.76 | | |
| Cancer (yes) | 1774 | 15.59 | 1721 | 14.93 | 2 | 0.173 |
| Cancer (no) | 9607 | 84.41 | 9805 | 85.07 | | |
| Alzheimer's disease (yes) | 26 | 0.23 | 25 | 0.22 | 0.002 | 0.962 |
| Alzheimer's disease (no) | 11357 | 99.77 | 11510 | 99.78 | | |
| Smoking status (current) | 941 | 8.29 | 975 | 8.49 | 134 | 7.65e-30 |
| Smoking status (former) | 7309 | 64.42 | 6589 | 57.38 | | |
| Smoking status (never) | 3095 | 27.28 | 3920 | 34.13 | | |
| Medication burden (median) | 0-5 | 0 | 0-6 | 1 | 137 | 1.13e-31 |

| | | | | | | |
|---|---|---|---|---|---|---|
| Body mass index (kg/m$^2$) | 13.55-59.95 | 28.34 (4.71) | 12.90-69.65 | 27.81 (5.97) | 7.4 | 1.58e-13 |
| Menopause (yes) | - | - | 7741 | 80.46 | - | - |
| Menopause (no) | - | - | 1880 | 19.54 | - | - |
| Age at menopause (years) | - | - | 11.00-75.00 | 49.96 (5.15) | - | - |
| Number of live births | - | - | 0-35 | 2.316 (1.46) | - | - |
| Ever used hormonal contraceptives (yes) | - | - | 8544 | 79.81 | - | - |
| Ever used hormonal contraceptives (no) | - | - | 2161 | 20.19 | - | - |
| Ever used hormone-replacement therapy (yes) | - | - | 4430 | 38.51 | - | - |
| Ever used hormone-replacement therapy (no) | - | - | 7073 | 61.49 | - | - |

Welch's two sample t-test was used to compare means of continuous variables and Pearson's Chi-squared test with Yates' continuity correction for counts of categorical variables. Kruskal-Wallis rank sum test was used for medication burden specifically due to the non-normal distribution of count data. All *p*-values are two-sided.

**Table 4:** Clinically acceptable ranges for circulating biomarkers collected from Mayo Clinic guidelines.

| Measure | Source | Sex | Lower bound | Upper bound | Units |
|---|---|---|---|---|---|
| White blood cell (leukocyte) count | Complete blood count | Both | 3.4 | 9.6 | $10^9$/L |
| Red blood cell (erythrocyte) count | Complete blood count | Male | 4.35 | 5.65 | $10^{12}$/L |
| Red blood cell (erythrocyte) count | Complete blood count | Female | 3.92 | 5.13 | $10^{12}$/L |
| Hemoglobin concentration | Complete blood count | Male | 13.2 | 16.6 | g/dL |
| Hemoglobin concentration | Complete blood count | Female | 11.6 | 15 | g/dL |
| Hematocrit percentage | Complete blood count | Male | 38.3 | 48.6 | % |
| Hematocrit percentage | Complete blood count | Female | 35.5 | 44.9 | % |
| Mean corpuscular volume | Complete blood count | Male | 78.2 | 97.9 | fL |
| Mean corpuscular volume | Complete blood count | Female | 78.2 | 97.93 | fL |
| Mean corpuscular hemoglobin* | Complete blood count | Both | 27 | 31 | pg/cell |
| Mean corpuscular hemoglobin concentration* | Complete blood count | Both | 32 | 36 | g/dL |
| Red blood cell (erythrocyte) distribution width | Complete blood count | Male | 11.8 | 14.5 | % |
| Red blood cell (erythrocyte) distribution width | Complete blood count | Female | 12.2 | 16.1 | % |
| Platelet count | Complete blood count | Male | 135 | 317 | $10^9$/L |
| Platelet (thrombocyte) count | Complete blood count | Female | 157 | 371 | $10^9$/L |
| Mean platelet (thrombocyte) volume** | Complete blood count | Both | 7 | 9 | fL |
| Lymphocyte count | Blood differential | Both | 0.95 | 3.07 | $10^9$/L |
| Monocyte count | Blood differential | Both | 0.26 | 0.81 | $10^9$/L |
| Granulocyte count*** | Blood differential | Both | 1.60 | 7.01 | $10^9$/L |
| Neutrophil count | Blood differential | Both | 1.56 | 6.45 | $10^9$/L |
| Eosinophil count | Blood differential | Both | 0.03 | 0.48 | $10^9$/L |
| Basophil count | Blood differential | Both | 0.01 | 0.08 | $10^9$/L |
| Albumin | Metabolic panel | Both | 3.5 | 5 | g/dL |
| Alanine aminotransferase | Metabolic panel | Male | 7 | 55 | U/L |
| Alanine aminotransferase | Metabolic panel | Female | 7 | 45 | U/L |
| Cholesterol**** | Lipid panel | Both | - | 200 | mg/dL |
| Creatinine***** | Metabolic panel | Male | 0.74 | 1.35 | mg/dL |
| Creatinine***** | Metabolic panel | Female | 0.59 | 1.04 | mg/dL |
| C-reactive protein | - | Both | - | 5 | mg/L |
| Glycated hemoglobin (HbA1c)****** | - | Both | 4 | 6.5 | % |
| HDL cholesterol**** | Lipid panel | Male | 40 | - | mg/dL |
| HDL cholesterol**** | Lipid panel | Female | 50 | - | mg/dL |
| LDL direct**** | Lipid panel | Both | - | 130 | mg/dL |
| Triglyceride**** | Lipid panel | Both | - | 150 | mg/dL |
| Vitamin D | - | Both | 10 | 80 | ng/mL |

**Note:** Clinical guidelines were identified from the Mayo Clinic unless otherwise noted
*Clinical guidelines identified from Mount Sinai Health System
**Clinical guidelines identified from the Clevland Clinic
***Clinical guidelines for total granulocyte count is a sum of the three cellular subtype cutoffs
****Values were converted to mmol/L based on this source: https://www.mayoclinic.org/diseases-conditions/high-blood-cholesterol/diagnosis-treatment/drc-20350806

*****Values were converted to µmol/L based on this source: https://www.mayoclinic.org/tests-procedures/creatinine-test/about/pac-20384646
******Values were converted to mmol/mol based on International Federation of Clinical Chemistry guidelines

**Table 5:** Box-Cox lambdas used to normalize the outcome measures in this study.

| Measure | UK Biobank | CLSA |
|---|---|---|
| White blood cell (leukocyte) count | 0.66 | 0.45 |
| Lymphocyte count | 0.34 | 0.38 |
| Monocyte count | 0.00 | -1.00 |
| Granulocyte count | - | 0.43 |
| Neutrophil count | 0.60 | - |
| Eosinophil count | 0.10 | - |
| Basophil count | 0.17 | - |
| Red blood cell (erythrocyte) count | -0.77 | -0.45 |
| Hemoglobin concentration | 0.63 | 0.61 |
| Hematocrit percentage | -0.26 | 0.74 |
| Mean corpuscular volume | 2.00 | 2.00 |
| Mean corpuscular hemoglobin | 2.00 | 2.00 |
| Mean corpuscular hemoglobin concentration | 2.00 | -1.00 |
| Red blood cell (erythrocyte) distribution width | -1.00 | -1.00 |
| Platelet (thrombocyte) count | 0.52 | -0.12 |
| Mean platelet (thrombocyte) volume | 2.00 | 1.57 |
| Albumin | 2.00 | -0.44 |
| Alanine aminotransferase | -0.13 | -0.10 |
| Creatinine | -0.53 | -0.10 |
| Cholesterol | 2.00 | 2.00 |
| HDL cholesterol | -0.54 | -0.58 |
| LDL direct | 2.00 | 1.55 |
| Glycated hemoglobin (HbA1c) | 0.94 | 1.03 |
| Triglyceride | 0.61 | -1.00 |
| Vitamin D | 0.81 | 1.76 |
| C-reactive protein | 0.15 | -0.03 |

**Table 6:** Likelihood ratio tests between nested models of increasing complexity in the UK Biobank.

| Outcome | Model | Degrees of Freedom | Likelihood Ratio Chi-squared | Sig. |
|---|---|---|---|---|
| White Blood Cell | 2 vs. 1 | 2 | 649.80 | * |
| | 3 vs. 2 | 3 | 1244.95 | * |
| | 4 vs. 2 | 3 | 13.18 | * |
| | 5 vs. 2 | 1 | 3.28 | NS |
| | 6 vs. 3 | 3 | 13.40 | * |
| | 7 vs. 3 | 1 | 4.27 | * |
| | 8 vs. 6 | 1 | 4.91 | * |
| | 8 vs. 7 | 3 | 14.03 | * |
| | 9 vs. 8 | 3 | 3.61 | NS |
| Neutrophil | 2 vs. 1 | 2 | 1055.38 | * |
| | 3 vs. 2 | 3 | 2492.40 | * |
| | 4 vs. 2 | 3 | 5.13 | NS |
| | 5 vs. 2 | 1 | 3.18 | NS |
| | 6 vs. 3 | 3 | 5.35 | NS |
| | 7 vs. 3 | 1 | 4.82 | * |
| | 8 vs. 6 | 1 | 5.20 | * |
| | 8 vs. 7 | 3 | 5.73 | NS |
| | 9 vs. 8 | 3 | 5.93 | NS |
| Lymphocyte | 2 vs. 1 | 2 | 87.12 | * |
| | 3 vs. 2 | 3 | 457.53 | * |
| | 4 vs. 2 | 3 | 4.32 | NS |
| | 5 vs. 2 | 1 | 0.07 | NS |
| | 6 vs. 3 | 3 | 4.33 | NS |
| | 7 vs. 3 | 1 | 0.19 | NS |
| | 8 vs. 6 | 1 | 0.15 | NS |
| | 8 vs. 7 | 3 | 4.29 | NS |
| | 9 vs. 8 | 3 | 1.17 | NS |
| C-reactive Protein | 2 vs. 1 | 2 | 32.80 | * |
| | 3 vs. 2 | 3 | 75.20 | * |
| | 4 vs. 2 | 3 | 17.64 | * |
| | 5 vs. 2 | 1 | 0.48 | NS |
| | 6 vs. 3 | 3 | 17.46 | * |
| | 7 vs. 3 | 1 | 0.38 | NS |
| | 8 vs. 6 | 1 | 0.62 | NS |
| | 8 vs. 7 | 3 | 17.70 | * |
| | 9 vs. 8 | 3 | 2.21 | NS |
| Triglyceride | 2 vs. 1 | 2 | 185.87 | * |
| | 3 vs. 2 | 3 | 1487.72 | * |
| | 4 vs. 2 | 3 | 6.05 | NS |
| | 5 vs. 2 | 1 | 9.03 | * |
| | 6 vs. 3 | 3 | 7.47 | NS |
| | 7 vs. 3 | 1 | 10.73 | * |
| | 8 vs. 6 | 1 | 9.56 | * |
| | 8 vs. 7 | 3 | 6.29 | NS |
| | 9 vs. 8 | 3 | 0.54 | NS |

1. outcome ~ PRS + sex + age + covariates
2. outcome ~ PRS + sex + rcs(age, 4) + covariates
3. outcome ~ PRS + sex + rcs(age, 4) + sex:rcs(age, 4) + covariates
4. outcome ~ PRS + sex + rcs(age, 4) + PRS:rcs(age,4) + covariates
5. outcome ~ PRS + sex + rcs(age, 4) + PRS:sex + covariates
6. outcome ~ PRS + sex + rcs(age, 4) + sex:rcs(age, 4) + PRS:rcs(age, 4) + covariates
7. outcome ~ PRS + sex + rcs(age, 4) + sex:rcs(age, 4) + PRS:sex + covariates
8. outcome ~ PRS + sex + rcs(age, 4) + sex:rcs(age, 4) + PRS:rcs(age, 4) + PRS:sex + covariates
9. outcome ~ PRS + sex + rcs(age, 4) + sex:rcs(age, 4) + PRS:rcs(age, 4) + PRS:sex + PRS:rcs(age, 4):sex + covariates

Covariates = medication burden + BMI + smoking status + anxiety + depression + stroke + cancer + Alzheimer's disease + data collection site + fasting time

Note: Sig. is denoted as * if $p<0.05$ and NS if $p>0.05$.

**Table 7:** Likelihood ratio tests between nested models of increasing complexity in the CLSA.

| Outcome | Model | Degrees of Freedom | Likelihood Ratio Chi-squared | Sig. |
|---|---|---|---|---|
| White Blood Cell | 2 vs. 1 | 2 | 111.68 | * |
| | 3 vs. 2 | 3 | 57.37 | * |
| | 4 vs. 2 | 3 | 2.37 | NS |
| | 5 vs. 2 | 1 | 0.56 | NS |
| | 6 vs. 3 | 3 | 2.30 | NS |
| | 7 vs. 3 | 1 | 0.68 | NS |
| | 8 vs. 6 | 1 | 0.74 | NS |
| | 8 vs. 7 | 3 | 2.36 | NS |
| | 9 vs. 8 | 3 | 1.05 | NS |
| Granulocyte | 2 vs. 1 | 2 | 158.11 | * |
| | 3 vs. 2 | 3 | 107.62 | * |
| | 4 vs. 2 | 3 | 5.87 | NS |
| | 5 vs. 2 | 1 | 0.03 | NS |
| | 6 vs. 3 | 3 | 5.49 | NS |
| | 7 vs. 3 | 1 | 0.00 | NS |
| | 8 vs. 6 | 1 | 0.00 | NS |
| | 8 vs. 7 | 3 | 5.49 | NS |
| | 9 vs. 8 | 3 | 0.67 | NS |
| Lymphocyte | 2 vs. 1 | 2 | 1.33 | NS |
| | 3 vs. 2 | 3 | 22.40 | * |
| | 4 vs. 2 | 3 | 0.77 | NS |
| | 5 vs. 2 | 1 | 1.75 | NS |
| | 6 vs. 3 | 3 | 0.76 | NS |
| | 7 vs. 3 | 1 | 1.59 | NS |
| | 8 vs. 6 | 1 | 1.61 | NS |
| | 8 vs. 7 | 3 | 0.77 | NS |
| | 9 vs. 8 | 3 | 3.98 | NS |
| C-reactive Protein | 2 vs. 1 | 2 | 4.04 | NS |
| | 3 vs. 2 | 3 | 4.29 | NS |
| | 4 vs. 2 | 3 | 0.92 | NS |
| | 5 vs. 2 | 1 | 1.28 | NS |
| | 6 vs. 3 | 3 | 0.92 | NS |
| | 7 vs. 3 | 1 | 1.22 | NS |
| | 8 vs. 6 | 1 | 1.22 | NS |
| | 8 vs. 7 | 3 | 0.92 | NS |
| | 9 vs. 8 | 3 | 6.82 | NS |
| Triglyceride | 2 vs. 1 | 2 | 26.69 | * |
| | 3 vs. 2 | 3 | 50.63 | * |
| | 4 vs. 2 | 3 | 0.45 | NS |
| | 5 vs. 2 | 1 | 0.55 | NS |
| | 6 vs. 3 | 3 | 0.51 | NS |
| | 7 vs. 3 | 1 | 0.33 | NS |
| | 8 vs. 6 | 1 | 0.41 | NS |
| | 8 vs. 7 | 3 | 0.58 | NS |
| | 9 vs. 8 | 3 | 1.04 | NS |

1. outcome ~ PRS + sex + age + covariates
2. outcome ~ PRS + sex + rcs(age, 4) + covariates
3. outcome ~ PRS + sex + rcs(age, 4) + sex:rcs(age, 4) + covariates
4. outcome ~ PRS + sex + rcs(age, 4) + PRS:rcs(age,4) + covariates
5. outcome ~ PRS + sex + rcs(age, 4) + PRS:sex + covariates
6. outcome ~ PRS + sex + rcs(age, 4) + sex:rcs(age, 4) + PRS:rcs(age, 4) + covariates
7. outcome ~ PRS + sex + rcs(age, 4) + sex:rcs(age, 4) + PRS:sex + covariates
8. outcome ~ PRS + sex + rcs(age, 4) + sex:rcs(age, 4) + PRS:rcs(age, 4) + PRS:sex + covariates
9. outcome ~ PRS + sex + rcs(age, 4) + sex:rcs(age, 4) + PRS:rcs(age, 4) + PRS:sex + PRS:rcs(age, 4):sex + covariates

Covariates = medication burden + BMI + smoking status + anxiety + depression + stroke + cancer + Alzheimer's disease

Note: Sig. is denoted as * if $p<0.05$ and NS if $p>0.05$.

**Table 8:** Mediation models testing bidirectional mediation through a depression diagnosis in the CLSA.

| Predictor | → | Mediator | → | Outcome | Mediated | Direct | Proportion | Total |
|---|---|---|---|---|---|---|---|---|
| PRS | → | MDD | → | WBC | $4.68 \times 10^{-4}$ (0.43) | 0.0276 | 0.0166 (0.43) | 0.0281 |
| PRS | → | WBC | → | MDD | $6.85 \times 10^{-5}$ (0.44) | 0.0303 | 0.0023 (0.44) | 0.0304 |
| PRS | → | MDD | → | Granulocyte | $-3.92 \times 10^{-4}$ (0.5) | 0.0260 | -0.0153 (0.50) | 0.0256 |
| PRS | → | Granulocyte | → | MDD | $2.61 \times 10^{-5}$ (0.73) | 0.0318 | $8.18 \times 10^{-4}$ (0.73) | 0.0319 |
| PRS | → | MDD | → | Lymphocyte | 0.0017 | 0.0253 | 0.0640 | 0.0270 |
| PRS | → | Lymphocyte | → | MDD | $2.36 \times 10^{-4}$ | 0.0305 | 0.0077 | 0.0308 |
| PRS | → | MDD | → | CRP | 0.0012 | 0.0242 | 0.0463 | 0.0254 |
| PRS | → | CRP | → | MDD | $1.77 \times 10^{-4}$ | 0.0296 | 0.0059 | 0.0298 |
| PRS | → | MDD | → | Triglyceride | 0.0010 (0.20) | 0.0232 | 0.0431 (0.20) | 0.0242 |
| PRS | → | Triglyceride | → | MDD | $1.19 \times 10^{-4}$ (0.14) | 0.0283 | 0.0042 (0.14) | 0.0284 |

**Depression diagnosis (MDD) as a mediator**
**Mediator model:** MDD ~ PRS + sex + rcs(age, 4) + sex:rcs(age, 4) + covariates
**Outcome model:** biomarker ~ PRS + MDD + sex + rcs(age, 4) + sex:rcs(age, 4) + covariates

**Biomarker as a mediator**
**Mediator model:** biomarker ~ PRS + sex + rcs(age, 4) sex:rcs(age, 4) + covariates
**Outcome model:** MDD ~ PRS + biomarker + sex + rcs(age, 4) + sex:rcs(age, 4) + covariates

**Covariates** = BMI + smoking status + stroke + cancer + Alzheimer's disease
**Note:** Effects are reported from causal mediation analyses using nonparametric bootstrapped confidence intervals (1000 iterations). Effects are significant ($p$-value < 0.05) unless otherwise reported as "effect ($p$-value)"

**Table 9:** Mediation models testing bidirectional mediation through self-harm behaviors in the UK Biobank.

| Predictor | → | Mediator | → | Outcome | Mediated | Direct | Proportion | Total |
|---|---|---|---|---|---|---|---|---|
| PRS | → | Q1 | → | WBC | $4.35x10^{-4}$ | 0.0150 | 0.0282 | 0.0154 |
| PRS | → | WBC | → | Q1 | $4.63x10^{-5}$ | 0.0311 | 0.0015 | 0.0311 |
| PRS | → | Q2 | → | WBC | $1.33x10^{-4}$ (0.45) | 0.0153 | 0.0086 (0.45) | 0.0155 |
| PRS | → | WBC | → | Q2 | $1.34x10^{-5}$ (0.39) | 0.0204 | $6.54x10^{-4}$ (0.39) | 0.0204 |
| PRS | → | Q3 | → | WBC | $4.56x10^{-5}$ (0.67) | 0.0154 | 0.0030 (0.67) | 0.0154 |
| PRS | → | WBC | → | Q3 | $4.92x10^{-6}$ (0.61) | 0.0093 | $5.28x10^{-4}$ (0.61) | 0.0093 |
| PRS | → | Q1 | → | Neutrophil | $5.36x10^{-4}$ | 0.0113 | 0.0454 | 0.0118 |
| PRS | → | Neutrophil | → | Q1 | $4.21x10^{-5}$ | 0.0313 | 0.0013 | 0.0313 |
| PRS | → | Q2 | → | Neutrophil | $2.82x10^{-4}$ (0.09) | 0.0115 | 0.0239 (0.09) | 0.0118 |
| PRS | → | Neutrophil | → | Q2 | $2.08x10^{-5}$ (0.08) | 0.0203 | 0.0010 (0.08) | 0.0203 |
| PRS | → | Q3 | → | Neutrophil | $4.67x10^{-6}$ (0.93) | 0.0120 | $3.89x10^{-4}$ (0.93) | 0.0120 |
| PRS | → | Neutrophil | → | Q3 | $1.15x10^{-6}$ (0.84) | 0.0094 | $1.22x10^{-4}$ (0.84) | 0.0094 |
| PRS | → | Q1 | → | Lymphocyte | $4.78x10^{-5}$ (0.82) | 0.0071 | 0.0067 (0.82) | 0.0071 |
| PRS | → | Lymphocyte | → | Q1 | $2.85x10^{-6}$ (0.73) | 0.0313 | $9.12x10^{-5}$ (0.73) | 0.0313 |
| PRS | → | Q2 | → | Lymphocyte | $7.09x10^{-5}$ (0.63) | 0.0072 | 0.0098 (0.64) | 0.0073 |
| PRS | → | Lymphocyte | → | Q2 | $3.90x10^{-6}$ (0.62) | 0.0207 | $1.89x10^{-4}$ (0.62) | 0.0207 |
| PRS | → | Q3 | → | Lymphocyte | $1.35x10^{-4}$ (0.25) | 0.0070 | 0.0191 (0.26) | 0.0071 |
| PRS | → | Lymphocyte | → | Q3 | $5.63x10^{-6}$ (0.23) | 0.0093 | $6.04x10^{-4}$ (0.23) | 0.0093 |
| PRS | → | Q1 | → | CRP | $1.53x10^{-4}$ (0.45) | 0.0078 | 0.0193 (0.45) | 0.0080 |
| PRS | → | CRP | → | Q1 | $9.60x10^{-6}$ (0.45) | 0.0307 | $3.13x10^{-4}$ (0.45) | 0.0307 |
| PRS | → | Q2 | → | CRP | $1.48x10^{-4}$ (0.41) | 0.0074 | 0.0196 (0.41) | 0.0075 |
| PRS | → | CRP | → | Q2 | $7.92x10^{-6}$ (0.37) | 0.0199 | $3.98x10^{-4}$ (0.37) | 0.0199 |
| PRS | → | Q3 | → | CRP | $-1.29x10^{-4}$ (0.3) | 0.0078 | -0.0168 (0.34) | 0.0077 |
| PRS | → | CRP | → | Q3 | $-4.44x10^{-6}$ (0.4) | 0.0092 | $-4.85x10^{-4}$ (0.4) | 0.0092 |
| PRS | → | Q1 | → | Triglyceride | $7.38x10^{-4}$ | 0.0111 | 0.0624 | 0.0118 |
| PRS | → | Triglyceride | → | Q1 | $6.65x10^{-5}$ | 0.0289 | 0.0023 | 0.0290 |
| PRS | → | Q2 | → | Triglyceride | $2.64x10^{-4}$ (0.13) | 0.0119 | 0.0216 (0.13) | 0.0122 |
| PRS | → | Triglyceride | → | Q2 | $2.23x10^{-5}$ (0.17) | 0.0183 | 0.0012 (0.17) | 0.0183 |
| PRS | → | Q3 | → | Triglyceride | $9.86x10^{-5}$ (0.40) | 0.0120 | 0.0082 (0.4) | 0.0121 |
| PRS | → | Triglyceride | → | Q3 | $5.69x10^{-6}$ (0.56) | 0.0082 | $6.92x10^{-4}$ (0.56) | 0.0082 |

**Self-harm behavior as a mediator**
**Mediator model:** Qi ~ PRS + sex + rcs(age, 4) + sex:rcs(age, 4) + covariates
**Outcome model:** biomarker ~ PRS + Qi + sex + rcs(age, 4) + sex:rcs(age, 4) + covariates

Q1 = "Ever thought that life was not worth living?"
Q2 = "Ever contemplated self-harm?"
Q3 = "Ever self-harmed?"

**Biomarker as a mediator**
**Mediator model:** biomarker ~ PRS + sex + rcs(age, 4) sex:rcs(age, 4) + covariates
**Outcome model:** Qi ~ PRS + biomarker + sex + rcs(age, 4) + sex:rcs(age, 4) + covariates

**Covariates** = BMI + smoking status + stroke + cancer + Alzheimer's disease
**Note:** Effects are reported from causal mediation analyses using nonparametric bootstrapped confidence intervals (1000 iterations). Effects are significant ($p$-value < 0.05) unless otherwise reported as "effect ($p$-value)"

### Figures

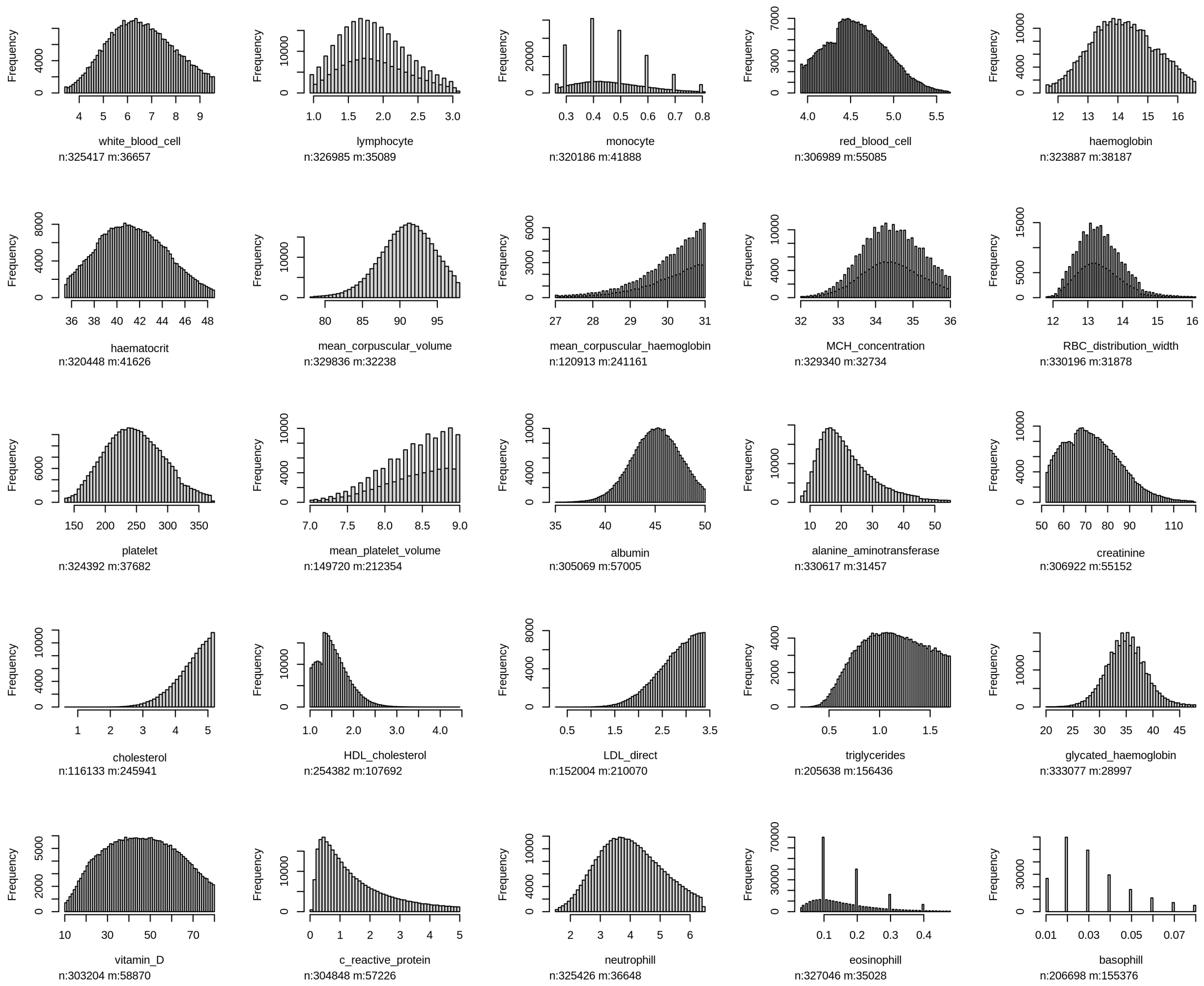


**Figure 1:** Histograms of the pruned measures in the UK Biobank ($n$ = sample size, $m$ = missing).

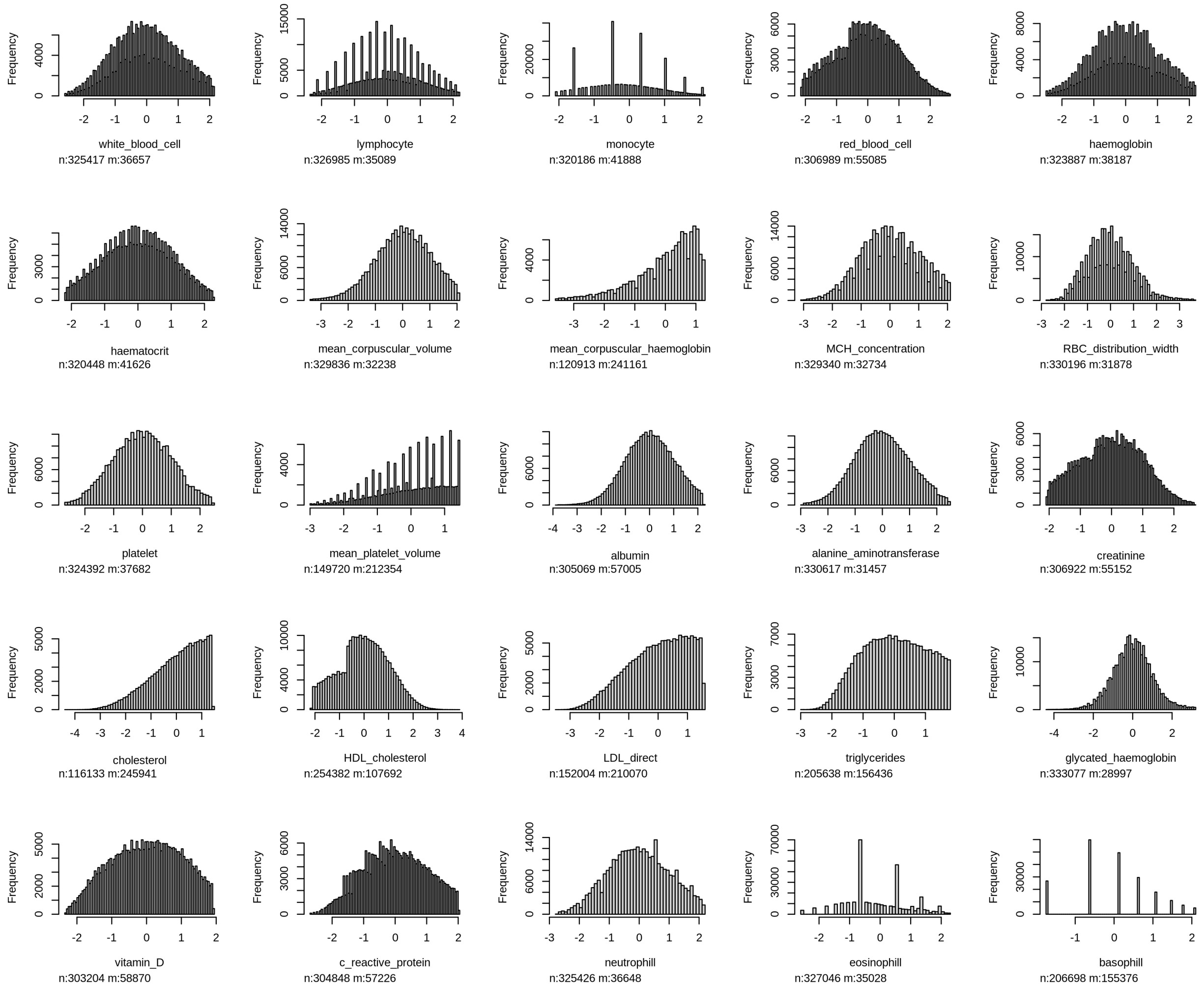


**Figure 2:** Histograms of the pruned and Box-Cox transformed measures in the UK Biobank ($n$ = sample size, $m$ = missing).

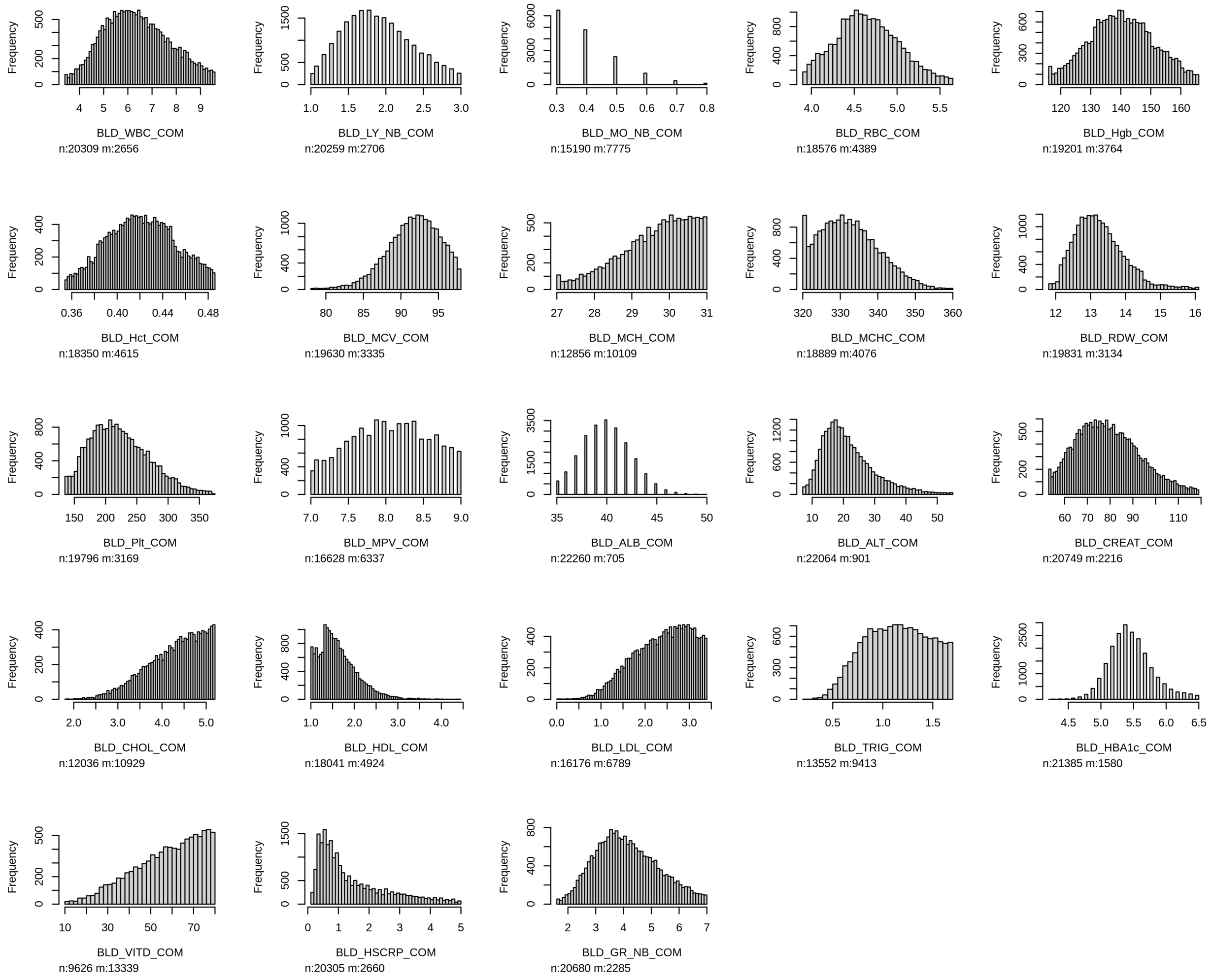


**Figure 3:** Histograms of the pruned measures in the CLSA (*n* = sample size, *m* = missing).

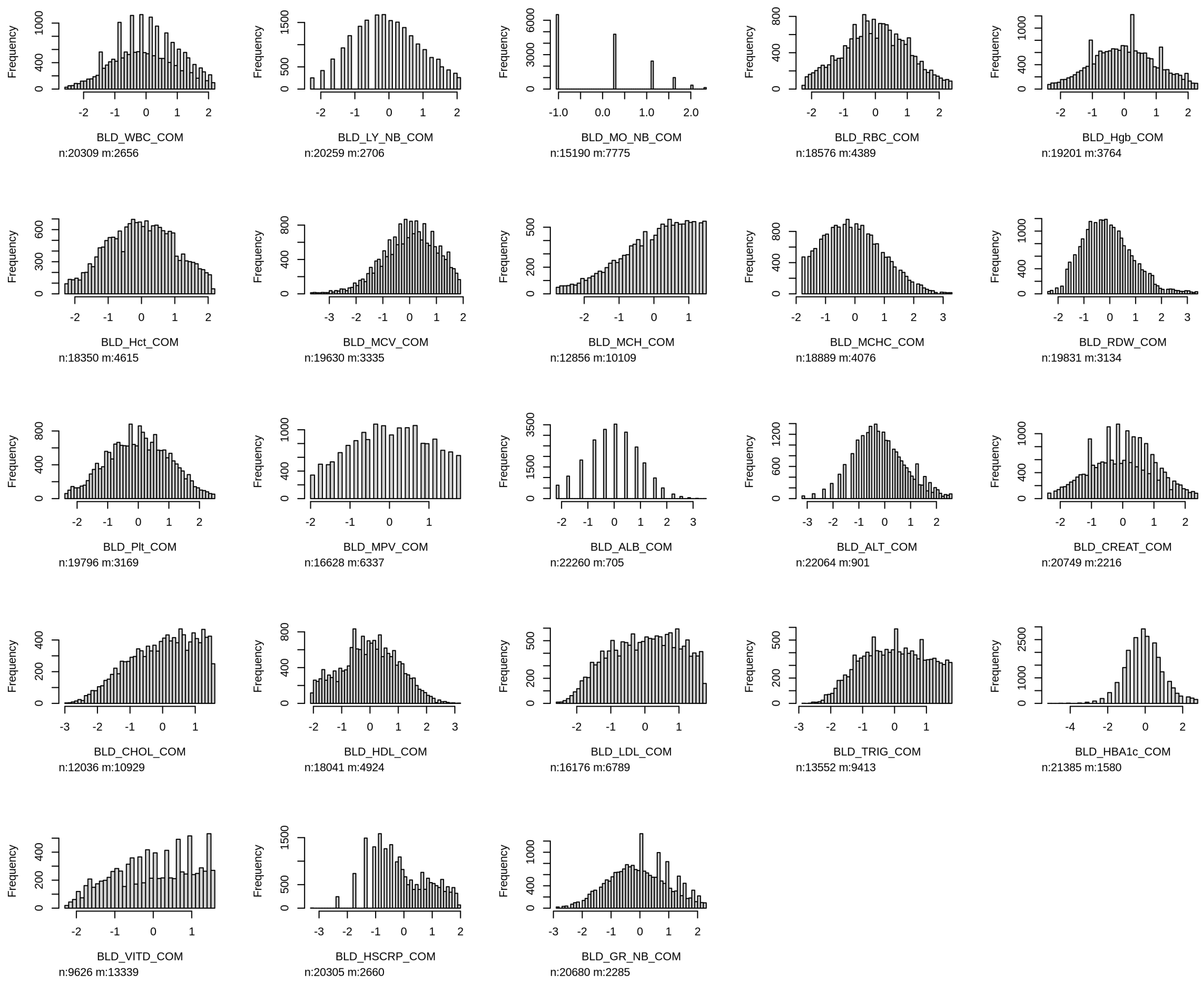


**Figure 4:** Histograms of the pruned and Box-Cox transformed measures in the CLSA (*n* = sample size, *m* = missing).
